# Persistent oxidative stress and inflammasome activation in CD14^high^CD16^-^ monocytes from COVID-19 patients

**DOI:** 10.1101/2021.09.13.21263292

**Authors:** Silvia Lucena Lage, Eduardo Pinheiro Amaral, Kerry L. Hilligan, Elizabeth Laidlaw, Adam Rupert, Sivaranjani Namasivayan, Joseph Rocco, Frances Galindo, Anela Kellogg, Princy Kumar, Rita Poon, Glenn W. Wortmann, John P. Shannon, Heather D. Hickman, Andrea Lisco, Maura Manion, Alan Sher, Irini Sereti

**Affiliations:** HIV Pathogenesis Section, Laboratory of Immunoregulation, National Institute of Allergy and Infectious Diseases, National Institutes of Health, Bethesda, MD, USA.; Immunobiology Section, Laboratory of Parasitic Diseases, National Institute of Allergy and Infectious Diseases, National Institutes of Health, Bethesda, MD, USA.; Immune Cell Biology Programme, Malaghan Institute of Medical Research, Wellington 6012, New Zealand.; AIDS Monitoring Laboratory, Frederick National Laboratory for Cancer Research, Leidos Biomedical Research, Inc, Frederick, MD, USA.; Clinical Monitoring Research Program, Frederick National Laboratory for Cancer Research, Leidos Biomedical Research, Inc, Frederick, MD, USA.; Division of Infectious Diseases and Travel Medicine at MedStar Georgetown University Hospital, Washington DC, USA.; Section of Infectious Diseases, MedStar Washington Hospital Center, Washington, DC, USA.; Viral Immunity and Pathogenesis Unit, Laboratory of Clinical Immunology and Microbiology, National Institutes of Allergy and Infectious Diseases, National Institutes of Health, Bethesda, MD, USA.

**Keywords:** COVID-19, NLRP3 inflammasome, Oxidative stress, Lipid peroxidation, CD14^high^CD16^-^ monocytes.

## Abstract

The poor outcome of the coronavirus disease-2019 (COVID-19), caused by SARS-CoV-2, is associated with systemic hyperinflammatory response and immunopathology. Although inflammasome and oxidative stress have independently been implicated in COVID-19, it is poorly understood whether these two pathways cooperatively contribute to disease severity. Herein, we found an enrichment of CD14^high^CD16^-^ monocytes displaying inflammasome activation evidenced by caspase-1/ASC-speck formation in severe COVID-19 patients when compared to mild ones and healthy controls, respectively. Those cells also showed aberrant levels of mitochondrial superoxide (MitoSOX) and lipid peroxidation, both hallmarks of the oxidative stress response, which strongly correlated with caspase-1 activity. In addition, we found that NLRP3 inflammasome-derived IL-1β secretion by SARS-CoV-2- exposed monocytes *in vitro* was partially dependent on lipid peroxidation. Importantly, altered inflammasome and stress responses persisted after short- term patient recovery. Collectively, our findings suggest oxidative stress/NLRP3 signaling pathway as a potential target for host-directed therapy to mitigate early COVID-19 hyperinflammation as well as its long-term outcomes.

## INTRODUCTION

Severe acute respiratory syndrome coronavirus 2 (SARS-CoV-2), the causative agent of coronavirus disease 2019 (COVID-19), emerged in late 2019 and rapidly spread worldwide, leading to approximately 3.6 million deaths to date (WHO, 2021). Although the majority of infected individuals are asymptomatic or experience mild disease, about 20% of patients with COVID-19 develop moderate/severe manifestations mainly characterized by hypoxia and extensive pneumonia (Wu et al., 2020). Patients occasionally progress to critical condition as a result of pathological complications related to acute respiratory distress syndrome (ARDS), endothelial dysfunction and coagulopathy, leading to multi-organ failure and consequently death (Ackermann et al., 2020; Bonaventura et al., 2021; Dandel, 2021; Gibson et al., 2020).

Although the factors involved in disease progression to severe forms remain poorly understood, an exacerbated inflammatory response has been implicated as a major cause of morbidity and mortality in patients with COVID-19 (Hojyo et al., 2020; Tang et al., 2020). Overall, elevated levels of circulating inflammatory markers including C-reactive protein (CRP), ferritin, D-dimer and a wide range of inflammatory cytokines/chemokines have been associated with poor disease outcome. Importantly, the pro-inflammatory cytokines IL-1β and IL-18 have both been implicated in lung injury and respiratory distress induced by SARS-CoV-2 (Conti et al., 2020; Meduri et al., 1995; Satis et al., 2021). Of note, mature IL-1β plays an important role in immune cell recruitment to the site of infection as well as in stimulating cells to produce and release a number of inflammatory mediators including IL-6 and CRP (Patton et al., 1995; Slaats et al., 2016). In addition, mature IL-18 has also been associated with hyperferritinemia and cytopenias, both pathological features observed in COVID-19 patients displaying severe disease (Agbuduwe and Basu, 2020; Perricone et al., 2020). IL-1β and IL-18 production rely on inflammasome activation upon sensing of pathogen/damage-associated molecular patterns (PAMPs/DAMPs) by distinct innate immune intracellular sensors (Martinon et al., 2002; Sharma and Kanneganti, 2016). Once activated, those sensors recruit the adapter molecule ASC (apoptosis-associated speck-like protein containing a caspase activating and recruitment domain, CARD) in order to activate pro-caspase-1, thus forming the inflammasome complex also known as ASC-speck (Lu et al., 2014). The enzymatic activity of caspase-1 is required for IL-1β and IL-18 processing and secretion as well as for the cleavage of the pore-forming protein gasdermin D (GSDMD), thus inducing a cell death process called pyroptosis (Broz et al., 2010; Shi et al., 2015).

Among the distinct cytosolic sensors able to form inflammasomes the molecule NLRP3 (NOD-, LRR- and pyrin domain-containing protein 3) has been implicated in COVID-19 (Abais et al., 2015; Freeman and Swartz, 2020; Rodrigues et al., 2021; Shah, 2020). Several stress-related cellular processes have been reported to induce NLRP3 activation in response to distinct PAMPs and DAMPs, such as K^+^ efflux (Munoz-Planillo et al., 2013), lysosomal disruption (Hornung et al., 2008), mitochondrial damage (Iyer et al., 2013; Zhou et al., 2011) and the generation of reactive oxygen species (ROS) (Martinon, 2010; Tschopp and Schroder, 2010). Since robust redox responses observed in hypoxic conditions during other pulmonary infections are commonly associated with exacerbated cytokine production and disease progression (Amaral et al., 2021; Erlich et al., 2020; Favier et al., 1994; Freeborn and Rockwell, 2021; Pohanka, 2013; Prabhakar et al., 2020), host oxidative stress has also been implicated in the pathogenesis of COVID-19 (Delgado-Roche and Mesta, 2020; Fakhri et al., 2020; Ntyonga-Pono, 2020).

Oxidative stress results from the imbalance of total oxidant status over antioxidant response (Bhat et al., 2012). Aberrant generation of ROS along with accumulation of toxic lipid peroxides can be detrimental to host cells and thus promote tissue damage. The inability of cells to detoxify biological membranes from lipid peroxidation is caused by a decrease in the enzymatic activity of glutathione peroxidase-4 (Gpx4) often due to reduced intracellular levels of its co-factor glutathione (GSH). Of note, a decrease in GSH levels has been reported in several comorbidities including type-2 diabetes (Goutzourelas et al., 2018; Lutchmansingh et al., 2018; Sekhar et al., 2011; Tan et al., 2012), obesity, cardiovascular disorders (Cavalca et al., 2009; Chaves et al., 2007; Shimizu et al., 2004), respiratory diseases (Fitzpatrick et al., 2012; Fitzpatrick et al., 2011; Fitzpatrick et al., 2009; Rahman and MacNee, 1999) cancer (Balendiran et al., 2004; Traverso et al., 2013) and hepatitis (Burgunder and Lauterburg, 1987; Shaw et al., 1983), most of which have been recognized as risk factors for the severe outcome of COVID-19. Moreover, it is possible that uncontrolled oxidative stress in the lung may form a positive feedback loop exacerbating NLRP3-inflammasome activation the production of pro-inflammatory cytokines such as IL-1β. In addition, the dysregulation of the cellular antioxidant response controlling lipid peroxidation may represent a potential host factor to be targeted to prevent aberrant inflammatory responses. However, the possible interplay of these two important inflammatory responses has not been formally investigated at the cellular level in COVID-19 patients.

Here, we assessed inflammasome and oxidative stress activation in circulating blood monocytes from COVID-19 patients and investigated whether these cellular pathways cooperatively contribute to COVID-19 disease severity and cytokine release syndrome. Our findings provide insights into the pathogenesis of COVID-19, both during acute infection and early recovery, suggesting inhibition of both NLRP3-inflammasome activation and lipid peroxidation as potential therapeutic interventions in COVID-19.

## RESULTS

### Significant changes in circulating monocyte subsets in COVID-19 patients

Dysregulated activation of the mononuclear phagocyte compartment leads to pathological inflammation and has been implicated in COVID-19 severity (Merad and Martin, 2020). We first investigated phenotypic changes in circulating blood monocytes from healthy control individuals (HCs) and COVID-19 patients experiencing mild or moderate-severe disease. Patients enrolled in this study were classified according to their oxygen requirement at the time of the research blood draw as (1) mild for patients requiring less than 4 Liters of oxygen (including no oxygen needed) (n=31; 23 inpatients and 8 outpatients, respectively), (2) moderate (n=4) for subjects who required up to 50% oxygen concentration, and (3) severe (n=12) for patients with a fraction of inspired oxygen (FiO2) over 50% or admitted to ICU care. We pooled patients displaying moderate and severe forms of the disease in the same group, which we named moderate-severe. Two patients remain in the critical care unit and one participant died by the time the study was completed. In addition, 14 patients were followed longitudinally after a short-term recovery period (approximately 52 days after infection onset), and 2 subjects were recruited after recovery. Clinical and demographic features of patients enrolled in the study are summarized in **Table 1**.

**TABLE 1.** COVID-19 participant characteristics (% or median values with IQR in parenthesis).

| **Characteristic** | **Mild** | **Moderate-**<br>**Severe** | **Recovered** |
| --- | --- | --- | --- |
| **N** | 31 | 16 | 16 |
| Age – y | 63 (50-67.5) | 62.5 (55.8-69.5) | 48.5 (37-57.5) |
| Female – no. (%) | 16 (52) | 7 (44) | 12 (75) |
| BMI (Kg/m2) | 31 (28.6-39.5) | 28.7 (24-36.4) | 31.5 (25.4-48.8) |
| **Race – no. (%)** |  |  |  |
| White | 13 (42) | 5 (31) | 9 (56) |
| African American | 14 (45) | 8 (50) | 3 (19) |
| Other | 4 (13) | 3 (19) | 4 (25) |
| *ANC (x10<sup>9</sup> cells/L) | 3.0 (2.4-4.6) | 7.3 (5.1-9.9) | 3.6 (3.1-4.9) |
| *ALC (x10<sup>9</sup> cells/L) | 1.3 (1.1-2.0) | 1.2 (0.9-1.8) | 1.9 (1.4-2.6) |
| Time from symptoms to lab draw (Days) | 9 (6-13.5) | 10 (8.8-11.5) | 52 (47.3-75.3) |
| **Comorbidities – no. (%)** |  |  |  |
| Hypertension | 18 (58) | 12 (75) | 8 (50) |
| Diabetes mellitus | 14 (45) | 6 (38) | 4 (25) |
| COPD/Asthma | 5 (16) | 5 (31) | 4 (25) |
| Renal Disease | 3 (10) | 2 (13) | 0 (0) |
| **Treatment – no. (%)** |  |  |  |
| Remdesivir | 6 (19) | 11 (69) | 4 (25) |
| Dexamethasone | 2 (6) | 4 (25) | 3 (19) |

\* ANC = Absolute neutrophil count, ALC = Absolute leukocyte count.

We observed that COVID-19 patients displayed a striking depletion of the patrolling CD14^low^CD16^+^ subset (Figure 1A-B) accompanied by enrichment of the CD14^high^CD16^-^ classical/inflammatory monocytes compared to HCs (Figure 1A and 1C). Moreover, CD14^high^CD16^-^ classical monocytes from COVID-19 patients showed downmodulation of HLA-DR and of the C–C chemokine receptor type 2 (CCR2) (Figure 1D and 1E, respectively). However, none of these features were associated with disease severity, since similar frequency of monocyte subsets and surface markers were observed when COVID-19 patients were grouped in mild and moderate-severe disease categories (Figure 1F-I). Collectively, these results demonstrate phenotypical and proportional changes in the peripheral monocytic compartment during SARS-CoV-2 infection consistent with previous reports (Wilk et al., 2020; Zhou et al., 2021)

**Figure 1.**
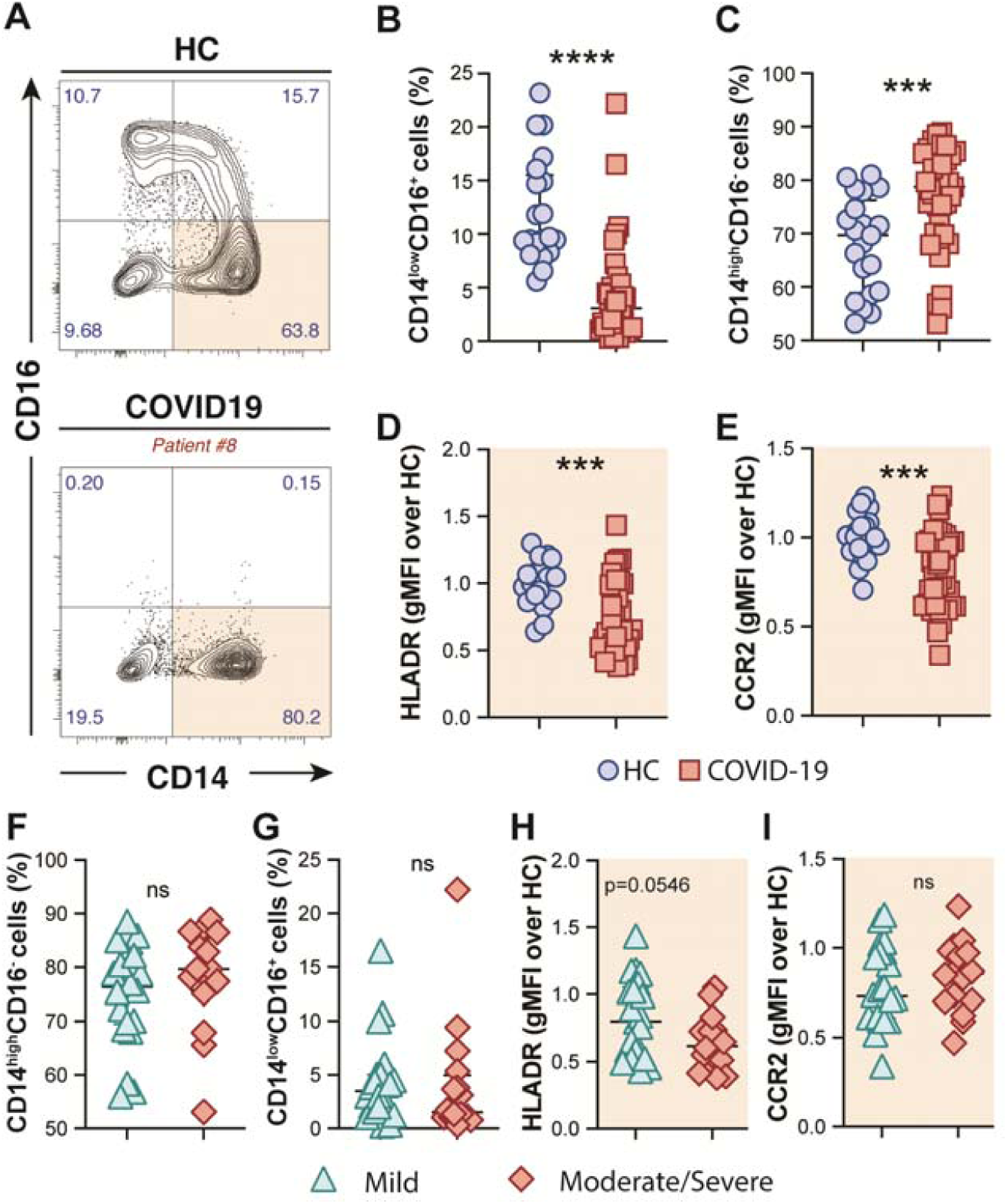
Phenotypic changes in circulating monocyte subsets during COVID-19. (**A**) Representative FACS plots showing the distribution of the monocyte subsets defined by CD14 and CD16 markers in COVID-19 patients. Percentage of CD14^low^CD16^+^ (**B**) and CD14^high^CD16^−^ (**C**) monocytes among circulating mononuclear myeloid cells (HLADR^+^CD2^−^CD3^−^CD19^−^CD20^−^CD56^−^CD66^−^) were compared between healthy controls (HC, n=21) and COVID-19 patients (n = 38). Data are presented as median with interquartile range. The geometric mean fluorescence intensity (gMFI) of HLA-DR (**D**) and CCR2 (**E**) expression on CD14^high^CD16^−^ monocytes was compared between HC (n=21) and COVID-19 patients (n=38). Percentage of CD14^high^CD16^-^ (**F**) and CD14^low^CD16^+^ (**G**) monocytes as well as MFI of HLA-DR (**H**) and CCR2 (**I**) expression on CD14^high^CD16^−^ monocytes is compared between COVID-19 patients experiencing mild symptoms (n=22) versus moderate to severe forms of COVID-19 (n=16). Data were analyzed using the Mann-Whitney test. ***P <0.001, ****P <0.0001; *ns*, not significant.

### Systemic inflammasome activation during COVID-19 is associated with disease severity

Systemic inflammation has been considered as a hallmark of COVID-19 severity (Hojyo et al., 2020). Accordingly, several molecular measurements associated with systemic inflammation including CRP, D-dimers, ferritin and IL-6 were elevated in plasma from COVID-19 patients in our cohort (Table 2 and Supplemental Figure 1). Additional analysis of these inflammatory markers failed to discriminate disease severity in this patient cohort (Supplemental Table 1). It is important to highlight that the COVID-19 patients were enrolled in the mild group based on their lower oxygen requirement (nasal cannula 4 Liters or less or no oxygen), which differs from other studies where only outpatients displaying no oxygen requirement were assigned as mild patients.

**TABLE 2.** Plasma biomarkers measurement in healthy controls (HC) versus COVID-19 patients (median values with IQR in parenthesis).

| **Biomarker**<br>**(pg/ml)** | **HC** | **COVID-19** | **p-value** |
| --- | --- | --- | --- |
| **N** | **13** | **29** | **NA** |
| **IFN-γ** | **5.34 (3.2-7.8)** | **9.17 (4.9-24.6)** | **0.06** |
| **IL-10** | **0.36 (0.24-0.65)** | **0.90 (0.60-2.33)** | **0.003** |
| **IL-12p70** | **0.40 (0.22-0.76)** | **0.31 (0.19-1.07)** | **0.94** |
| **IL-2** | **0.43 (0.29-0.47)** | **0.24 (0.13-0.40)** | **0.21** |
| **IL-6** | **0.48 (0.24-0.60)** | **6.07 (1.06-19.1)** | **<0.001** |
| **IL-8** | **2.41 (1.53-2.89)** | **4.36 (2.55-6.07)** | **0.03** |
| **TNF-α** | **1.50 (1.21-1.77)** | **3.02 (1.67-4.73)** | **0.002** |
| **IL-27** | **1410 (1095-1733)** | **2295 (1511-3770)** | **0.002** |
| **CRP (mg/L)** | **1.81 (1.55-7.45)** | **83.4 (9.30-187.9)** | **0.005** |
| **SAA (mg/L)** | **7.85 (3.98-9.96)** | **230.8 (14.9-404.8)** | **<0.001** |
| **sCD14 (mg/L)** | **1.36 (1.23-1.66)** | **1.75 (1.38-2.31)** | **0.05** |
| **IL-6Rα** | **32960 (30321-37174)** | **38853 (34737-44826)** | **0.02** |
| **D-dimer (ng/ml)** | **303 (214-579)** | **1452 (566-2502)** | **0.002** |
| **Ferritin (ng/ml)** | **101 (42.7-297)** | **1100 (173-1650)** | **0.007** |

\* **p-value** when Mann-Whitney was applied.

In addition to the above mentioned inflammatory markers, the inflammasome-derived cytokine IL-18 has also been implicated in COVID-19- associated pathological inflammation (Conti et al., 2020; Meduri et al., 1995; Satis et al., 2021). Accordingly, we observed higher plasma levels of IL-18 in our cohort of COVID-19 patients when compared to HCs (Figure 2A), suggesting systemic inflammasome activation in this group of patients. Since drastic depletion of the patrolling monocytes subset along with increased levels of classical/inflammatory CD14^high^CD16^-^ monocytes were observed in COVID-19 patients, we evaluated whether CD14^high^CD16^-^ cells represent a source of inflammasome activation during COVID-19. To do so, peripheral blood mononuclear cells (PBMCs) from HCs and patients were incubated with FLICA, an approach widely used to measure caspase-1/4/5 activity. We observed higher caspase-1/4/5 activity within the classical monocyte subset from COVID-19 patients when compared to HCs (Figure 2B), suggesting this cell subset contributes to IL-1β and IL-18 maturation during COVID-19.

**Figure 2.**
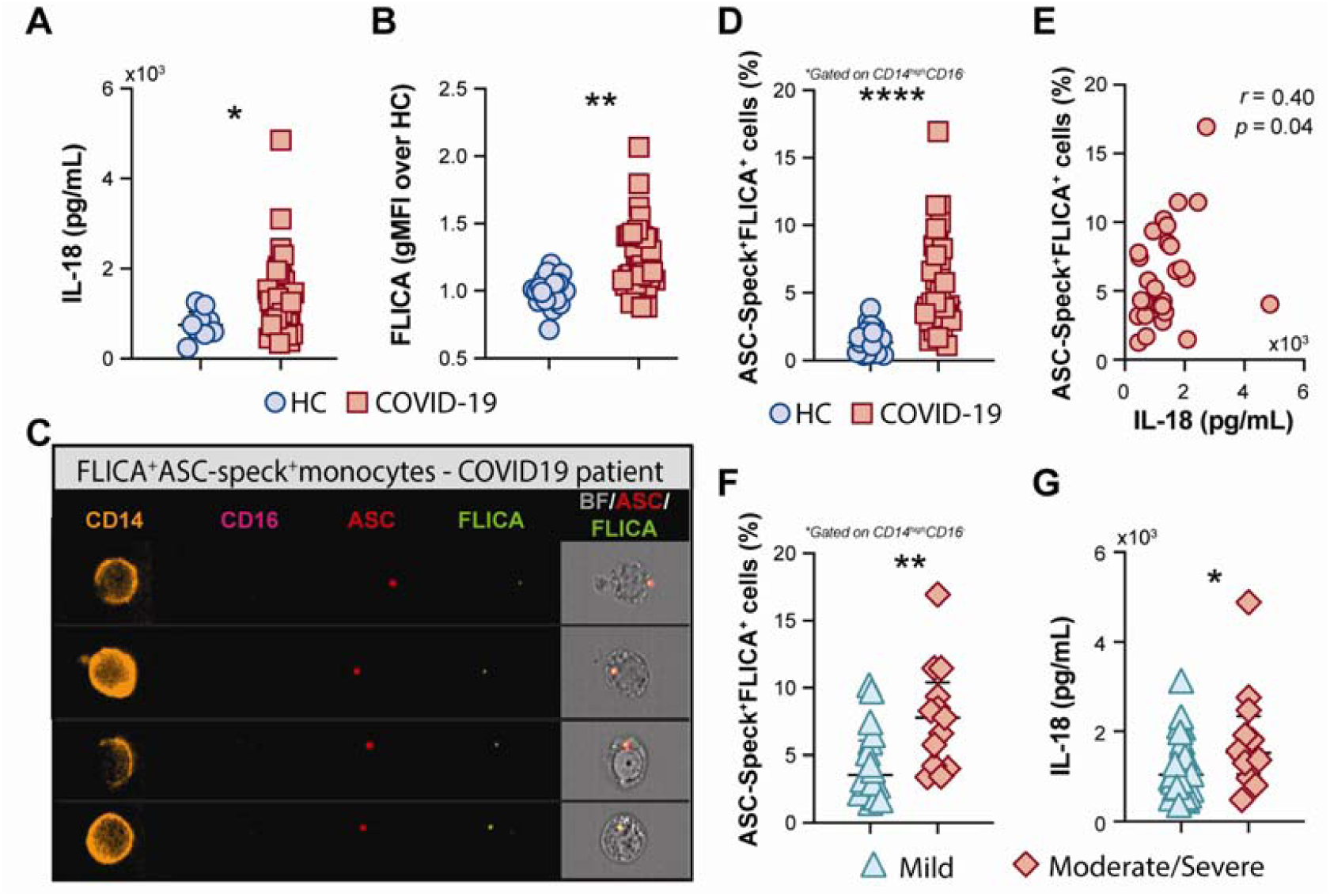
COVID-19 patients exhibit a high frequency of monocytes displaying FLICA^+^ASC-speck formation. (**A**) Plasma levels of IL-18 were compared between healthy control (HC) donors (n=9) and COVID-19 patients (n=39). Lines represent median values and interquartile ranges. **(B)** PBMCs were incubated with the fluorochrome inhibitor of caspase-1/4/5 (FLICA) and the geometric mean fluorescence intensity (gMFI) of FLICA within the CD14^high^CD16^−^ monocyte subset was compared between HC (n=21) and COVID-19 patients (n=38). Data are presented as median with interquartile range. (**C**) PBMCs from HC (n=24) and COVID-19 patients (n=32) were incubated with FLICA, stained for monocyte identification and intracellular ASC and acquired by using Imaging flow cytometry. Representative images showing respectively: CD14, CD16, ASC and FLICA fluorescence followed by a composite image containing brightfield (BF), and the fluorescence of ASC and FLICA merged, were selected from a COVID-19 patient. (**D**) The percentage of monocytes showing spontaneous FLICA^+^ASC-Speck formation was quantified after application of Modulation_Morphology (M11,Ch11)_11- ASC feature, followed by Bright Detail Similarity R3_MC_11-ASC_2-FLICA, inside the CD14^high^CD16^−^ monocyte gate, by using IDEAS software. Lines represent median values and interquartile ranges. (**E**) Spearman correlations between plasma levels of IL-18 and the percentage of ASC-speck^+^FLICA^+^ CD14^high^CD16^−^ monocytes in COVID-19 patient samples. Percentage of ASC- speck^+^FLICA^+^ cells (**F**) and plasma levels of IL-18 (**G**) were compared between COVID-19 patients experiencing mild symptoms (n=18 and n=28, respectively) and moderate to severe cases of COVID-19 (n=13 and n=12, respectively). Lines represent median values and interquartile ranges. Data were analyzed using the Mann-Whitney test. *P <0.05, **P <0.01, ****P <0.0001.

To directly evaluate inflammasome complex formation in CD14^high^CD16^-^ monocytes, we screened classical monocytes from HCs and COVID-19 patients for active ASC aggregates by imaging flow cytometry, following a previously described protocol (Lage et al., 2019). Briefly, PBMCs were pre-incubated with FLICA for caspase-1 detection followed by immunophenotyping and intracellular ASC staining. Using this approach, we identified monocytes with cytosolic ASC speck formation associated with active caspase-1/4/5, represented by FLICA^+^ASC-speck^+^ cells (Figure 2C, Merge panel). The summary data in Figure 2D demonstrate a substantially higher frequency of classical monocytes showing FLICA^+^ASC-speck^+^ formation in COVID-19 patients than HC subjects, which positively correlated with plasma IL-18 levels (Figure 2E). Importantly, in contrast to the other plasma inflammatory markers tested in this study (Supplemental Table 1), the percentage of monocytes with inflammasome complex formation and plasma levels of the downstream cytokine IL-18 were both significantly increased in the moderate-severe group than in patients with milder symptoms (Figure 2F and 2G, respectively), highlighting a role for inflammasomes in COVID-19 severity. Of note, inflammasome activation in circulating classical monocytes was not associated with age, sex, BMI, or race in our group of patients (Supplemental Figure 2).

Consistent with systemic inflammation, COVID-19-derived PBMCs but not HC cells spontaneously released IL-1β, as well as IL-6 and TNF-α *ex vivo* (Figure 3A, 3B and 3C, respectively). We next tested the effect of MCC950, a specific inhibitor of the NLRP3 inflammasome, and colchicine, an anti- inflammatory compound, on spontaneous cytokine release from COVID-19 patient cells. In addition to other anti-inflammatory effects, colchicine has been shown to inhibit microtubule-driven spatial arrangement of mitochondria, thus preventing full NLRP3 inflammasome activation (Misawa et al., 2013) and its use has been also reported to have beneficial effects in outpatients but not in hospitalized COVID-19 patients in recent clinical trials (Horby et al., 2021; Lopes et al., 2021; Tardif et al., 2021). Despite the promising clinical use of colchicine to prevent inflammasome activation, we did not observe any effect of compound on cytokine release by patient-derived PBMCs *in vitro* (Figure 3D-F). Conversely, selective inhibition of the inflammasome sensor NLRP3 by MCC950 specifically abolished IL-1β secretion *ex vivo* by PBMCs from COVID-19 patients without impacting IL-6 and TNF-α production (Figure 3D, 3E and 3F, respectively). Collectively, our findings support a role for NLRP3-mediated ASC-speck formation and caspase-1 activation in pathologic inflammation associated with moderate and severe forms of COVID-19.

**Figure 3.**
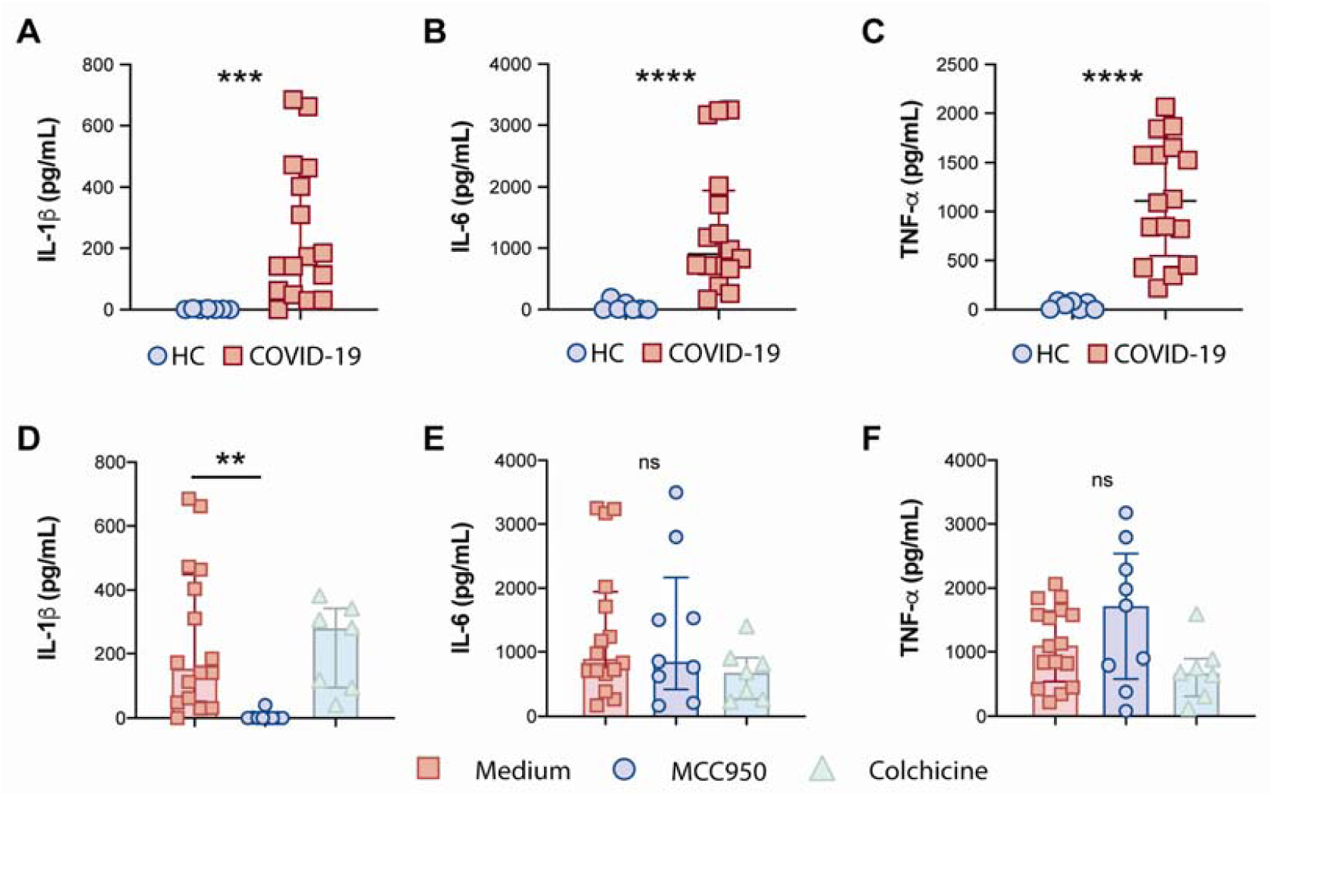
Effects of the NLRP3 inhibitor MCC950 and colchicine on cytokine secretion by COVID-19 PBMCs. IL-1β (**A**), IL-6 (**B**) and TNFα (**C**) levels from supernatants of healthy control (HC, n=7) or COVID-19 patients PBMCs (n=16) after a 4 hr incubation at 37C, were determined by multi- analyte flow assay kit. Lines represent median values and interquartile ranges. Alternatively, COVID-19 cells were pretreated with the NLRP3 inhibitor, MCC950 (10μM; n=9) or colchicine (10μM; n=7) during the incubation period, and IL-1β (**D**), IL-6 (**E**) and TNFα (**F**) levels were measured in the culture supernatants by multi-analyte flow assay kit. Numbers represent the median values and interquartile ranges. Data were analyzed using the Mann-Whitney test. **P <0.01, ***P <0.001, ****P <0.0001; *ns*, not significant.

### CD14^high^CD16^-^ monocytes from COVID-19 patients exhibit prominent oxidative stress activity

Oxidative stress has been suggested to be a critical host factor in driving immune response exacerbation in several infectious diseases (Amaral et al., 2021; Erlich et al., 2020; Favier et al., 1994; Freeborn and Rockwell, 2021; Pohanka, 2013). However, the induction of inflammatory oxidative stress responses in circulating monocytes obtained from COVID-19 patients has not been formally investigated. First, we sought to investigate the systemic oxidative response in plasma obtained from our cohort of COVID-19 patients by measuring the levels of ferritin heavy chain, catalase, total superoxide dismutase (SOD) activity, total antioxidant status, iron and lipid peroxidation (Figure 4A-F). Interestingly, elevated plasma levels of ferritin (Figure 4A), catalase (Figure 4B) superoxide dismutase (Figure 4C) and lipid peroxidation (Figure 4F) were found in COVID-19 patients compared to HC individuals whereas no significant difference was observed in the levels of total antioxidant response (Figure 4D) and iron (Figure 4F) in the same set of samples.

**Figure 4.**
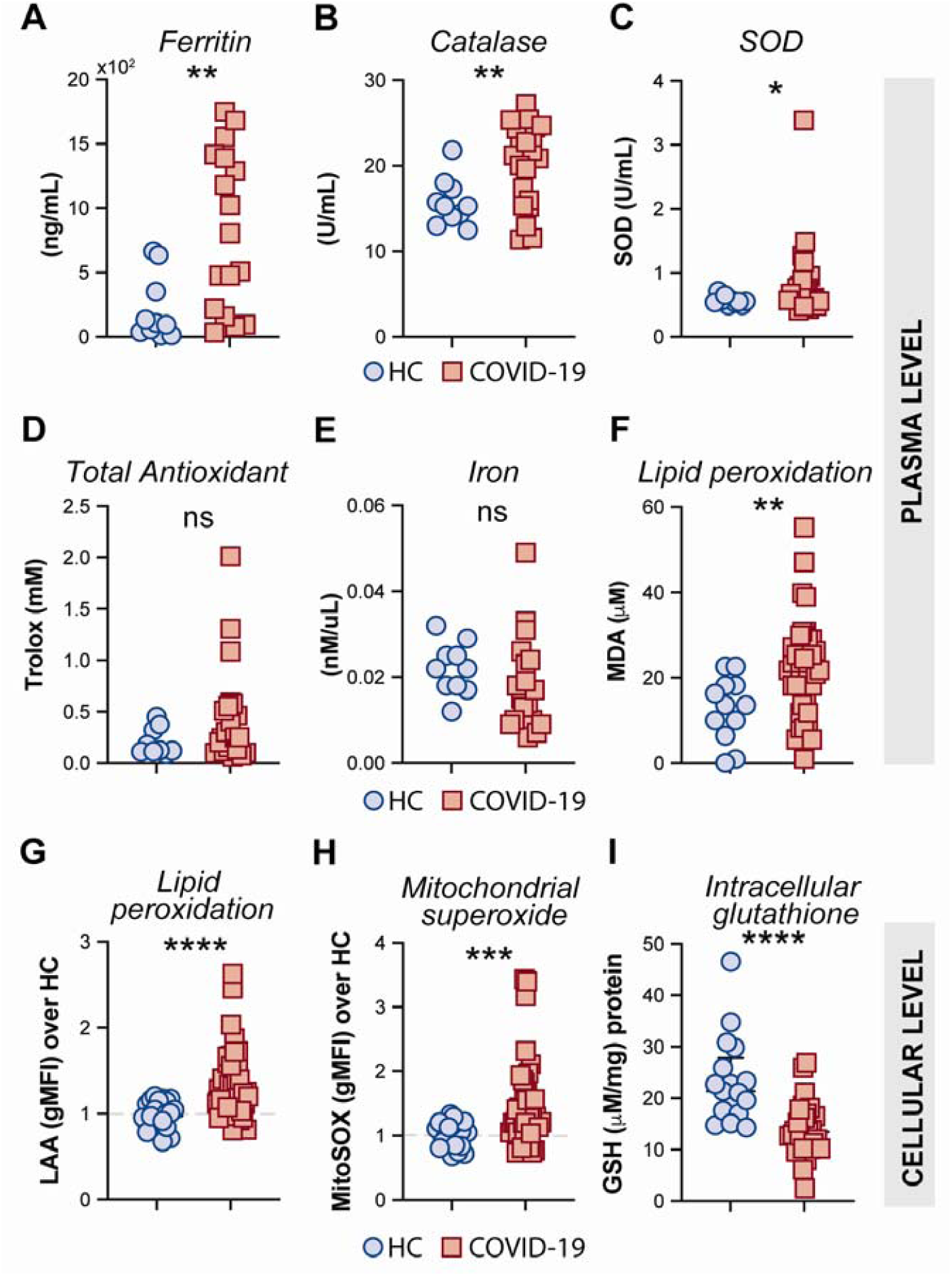
Measures of oxidative stress in COVID-19 patients. Plasma levels of ferritin (**A**), Catalase (**B**), Superoxide dismutase (SOD) (**C**), Trolox (**D**) Iron (**E**) and Lipid peroxidation (**F**) were compared between healthy control (HC) donors (n=10-12) and COVID-19 patients (n=20-40). Lipid peroxidation (LAA) (**G**) and mitochondrial superoxide (MitoSOX) (**H**) in CD14^high^CD16^−^ monocytes was measured by flow cytometry in PBMC samples from HC (n=22) and COVID-19 patients (n=40). Data are expressed as the geometric mean fluorescence intensity (gMFI) over average of HC. (**I**) Intracellular glutathione (GSH) levels in PBMC lysates from HC (n=17) and COVID-19 patients (n=27) were determined by analytic enzymatic assay as described in Methods. Data are presented as median values and interquartile ranges and analyzed using Mann-Whitney test. *P <0.05, **P <0.01, ***P <0.001, ****P <0.0001; *ns*, not significant.

Mitochondrial superoxide is an important intracellular source of reactive oxygen species and plays a critical role in promoting Fenton/Haber-Weiss reactions by releasing iron from intracellular ferritin complex, which in turn culminates in the generation of highly reactive hydroxyl radicals (Biemond et al., 1986; Biemond et al., 1984; Bou-Abdallah et al., 2018; Monteiro and Winterbourn, 1988; Williams et al., 1974). Elevated levels of hydroxy radicals in turn lead to abnormal generation and accumulation of toxic lipid peroxides affecting host cell function and viability. To carefully assess oxidative stress response in COVID-19 patients at the cellular level, we evaluated mitochondrial superoxide generation and lipid peroxidation in circulating CD14^high^CD16^-^ monocytes from COVID-19 patients (n=40) and HCs (n=23) by flow cytometric analysis. In addition, we used PBMC lysates from COVID-19 patients and HCs to investigate the host cellular antioxidant response by assessing intracellular levels of glutathione. We found elevated levels of mitochondrial superoxide along with aberrant lipid peroxidation in CD14^high^CD16^-^ monocytes from COVID-19 patients compared to HCs (Figure 4G and 4H, respectively). Consistent with these observations, a major reduction in intracellular glutathione levels was observed in PBMC lysates from COVID-19 patients compared to HCs (Figure 4I), suggesting an impaired intracellular host antioxidant response to control oxidative stress responses and ultimately lipid peroxidation.

### Exacerbated inflammasome activation is associated with mitochondrial dysfunction in COVID-19 patients

We next sought to evaluate associations between cellular inflammatory responses and oxidative stress in monocytes from COVID-19 patients and HCs. We found strong negative correlations between lipid peroxidation, mitochondrial superoxide and inflammasome activation and the frequency of CD14^low^CD16^+^ monocytes as well as CCR2 expression on CD14^high^CD16^-^ monocytes (Figure 5A). A highly significant negative correlation between glutathione levels and lipid peroxidation along with ASC-specks was also observed (Figure 5A), suggesting a possible link between cell stress responses and inflammasome activation during COVID-19.

**Figure 5.**
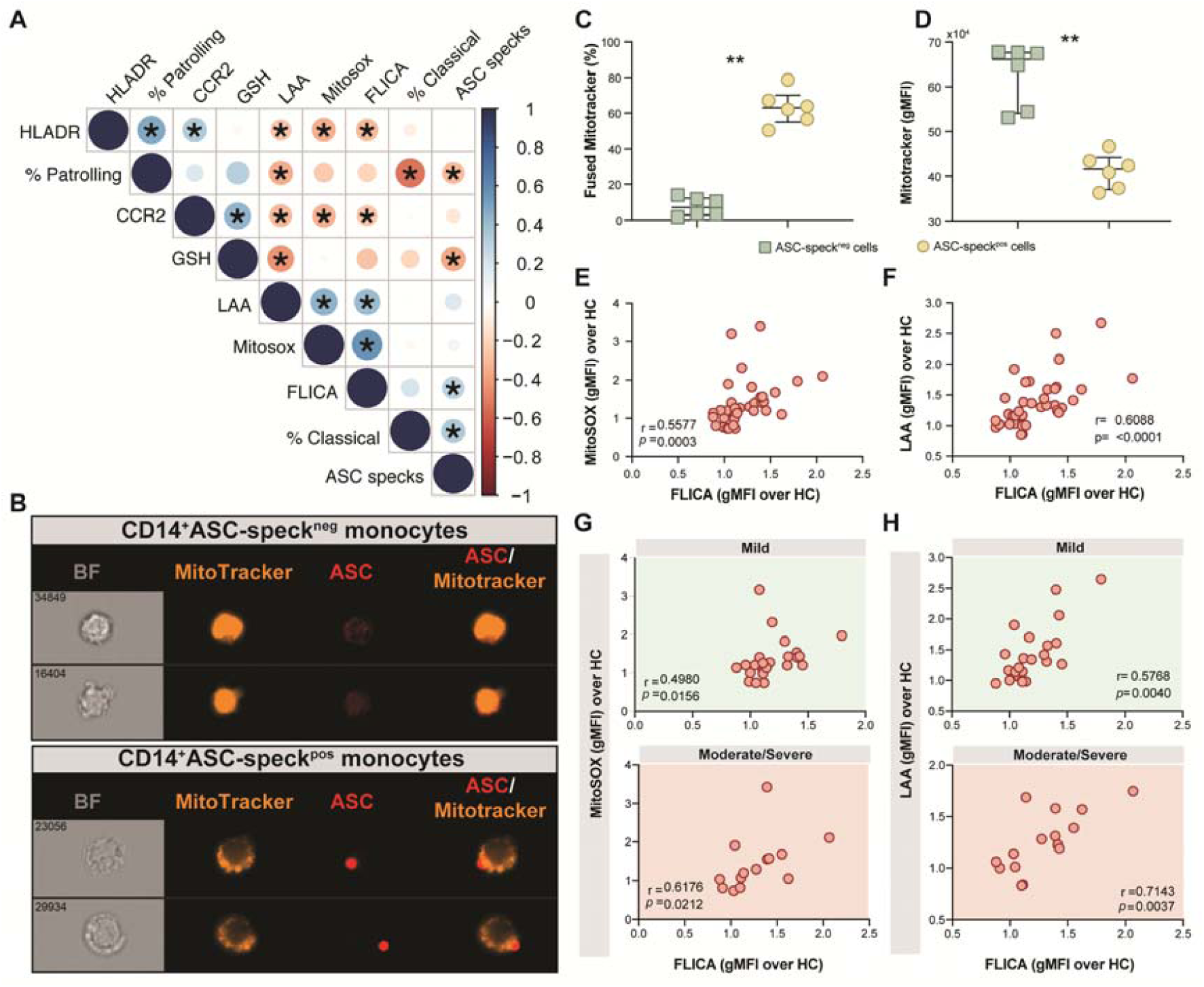
Oxidative stress and inflammasome responses are associated in COVID-19 patients. (**A**) Multi-parameter Spearman’s correlation analysis of PBMC samples from healthy control (HC) and COVID-19 patients (*P <0.05 or less). (**B**) PBMCs from COVID-19 patients (n=6) were incubated with Mitotracker Red, stained for intracellular ASC and acquired by means of Imaging flow cytometry. Representative images showing respectively: BF (brightfield), Mitotracker Red, ASC and a composite image of both Mitotracker Red and ASC markers merged, were selected from a CD14^+^ASC-speck^neg^ gate (top panel) and a CD14^+^ASC-speck^pos^ gate (bottom panel). (**C**) The percentage of monocytes showing the diffuse pattern for Mitotracker Red shown in B (top panel) *versus* the aggregated staining profile as shown in B (bottom panel) was determined after application of Major Axis Intensity_M04_Ch4 x Mean Pixel_M04_Ch4 features inside both CD14^+^ASC- speck^neg^ and CD14^+^ASC-speck^pos^ gates using IDEAS software. (**D**) Loss of mitochondrial membrane potential (ΔΨm) was determined by the change of Mitotracker Red gMFI staining on CD14^+^ASC-speck^neg^ and CD14^+^ASC-speck^pos^ subsets. Data are presented as median values and interquartile ranges and were analyzed using the Mann-Whitney test (**P < 0.01). (**E-F**) Spearman correlations between FLICA and MitoSOX (**E**) or FLICA and LAA (**F**) levels whithin CD14^high^CD16^−^ monocytes from COVID-19 patients. (**G-H**) Spearman correlations between FLICA and MitoSOX (**G**) or FLICA and LAA levels (**H**) on CD14^high^CD16^−^ monocytes obtained from COVID-19 patients grouped based on their disease severity in patients experiencing mild symptoms (top panel, green) or moderate to severe cases of COVID-19 (bottom panel, red). Each dot represents a correspondent value from each patient. *ns*, not significant.

Monocyte secretion of IL-1β IL-18 as a result of NLRP3 inflammasome activation requires mitochondrial-derived ROS and is commonly associated with an imbalance of cellular redox status (Martinon, 2010; Tschopp and Schroder, 2010). To assess the level of mitochondrial perturbation on monocytes isolated from COVID-19 patients, we incubated total PBMC with MitoTracker probe in order to evaluate mitochondrial membrane potential (ΔΨ in CD14 CD16 monocytes by imaging flow cytometry. This approach was also employed to address whether mitochondrial perturbation or dysfunction is associated with inflammasome assembly measured here by ASC-speck formation. CD14^high^CD16^-^ monocytes from COVID-19 patients were grouped according to their ASC-speck status in CD14^high^ASC-speck^neg^ (resting cells) and CD14^high^ASC-speck^pos^ cells and the loss of mitochondrial membrane potential was determined a decrease in MitoTracker MFI (Figure 5B). ASC-speck^pos^ monocytes showed an elevated frequency of cells displaying small puncta of MitoTracker staining compared to ASC-speck^neg^ cells (Figure 5C). In contrast, decreased MitoTracker gMFI was found in cells displaying ASC speck formation (Figure 5D), demonstrating loss of mitochondrial membrane potential and inflammasome activation in CD14^high^CD16^-^ monocytes.

Mitochondrial dysfunction is also linked with a metabolic shift from OXPHOS to glycolysis in inflammatory immune cells (Angajala et al., 2018; Monteiro et al., 2020). To investigate whether monocytes obtained from COVID-19 patients display this metabolic shift, we stained cells for Glut-1. Supporting our MitoTracker results, we found that CD14^high^CD16^-^ monocytes from COVID-19 patients showed elevated levels of Glut-1 (Supplemental Figure 3A), suggesting that these cells are undergoing intense cellular stress, reflecting by altered cell metabolism. Further, we found no difference in Glut-1 level when patients were grouped based on their disease severity status (Supplemental Figure 3B).

### Aberrant oxidative stress response strongly correlates with robust caspase-1 activation in COVID-19 patients

We have shown that COVID-19 patients display elevated caspase-1 activity and accumulation of lipid peroxides in circulating CD14^high^CD16^-^ monocytes. As the oxidative stress response expressed by lipid peroxidation also modulates NLRP3-inflammasome activation and the consequent release of mature IL-1β and IL-18 (Martinon, 2010; Tschopp and Schroder, 2010), we extended our analyses to explore the correlation between inflammasome activation and the oxidative stress response and their impact on disease severity.

We first investigated whether levels of mitochondrial superoxide in COVID-19 patients correlate with capase-1 activity. As shown in Figure 5E we observed a significant positive correlation between mitoSOX (gMFI) and FLICA (gMFI) in CD14^high^CD16^-^ monocytes from COVID-19 patients. In addition, we found a robust positive correlation between lipid peroxidation and active caspase-1 (Figure 5F). To test whether these parameters can reflect clinical disease severity, we stratified patients into mild or moderate-severe groups. We found a significant positive correlation of mitochondrial superoxide production and caspase-1 activity in monocytes from patients displaying mild and moderate/severe disease (Figure 5G). However, a stronger and more robust correlation was observed between lipid peroxidation and caspase-1 activity in patients in the moderate-severe group than those in the mild group (Figure 5H). Collectively, these findings reveal that CD14^high^CD16^-^ monocytes from COVID-19 patients display dysregulated oxidative stress responses and prominent inflammasome activation. Crosstalk between these two pathways may negatively impact the clinical course of SARS-CoV-2-infected subjects.

### IL-1**β** secretion by human monocytes in response to SARS-CoV-2 in vitro requires NLRP3 inflammasome activation and is partially dependent on lipid peroxidation

We next sought to investigate the crosstalk between the oxidative stress pathway and NLRP3-caspase-1 activation on circulating blood monocytes during COVID-19. To do so, we incubated column-purified monocytes isolated from healthy donors with SARS-CoV-2 USA-WA1/2020 at different multiplicities of infection (MOI) and measured IL-1β in supernatants after 24 hours as a result of inflammasome activation. As a negative control for active virus infection, monocytes were stimulated with heat-inactivated virus (HI-CoV-2) or viral media alone (Mock). First, to evaluate the presence of replicating virus in SARS-CoV-2-exposed monocyte cultures, we incubated VERO E6 cells, a cell lineage known to be sensitive to SARS-CoV-2 infection, with SARS-CoV-2-exposed monocyte lysates serially diluted. Although we failed to detect robust SARS-CoV-2 replication within human monocytes by using this method (Supplemental Figure 5), SARS-CoV-2 exposure to monocytes was sufficient to induce IL-1β secretion at MOI of 1, which was not seen in cultures incubated with HI-CoV-2 at the same MOI or with media only (mock) (Figure 6A).

**Figure 6.**
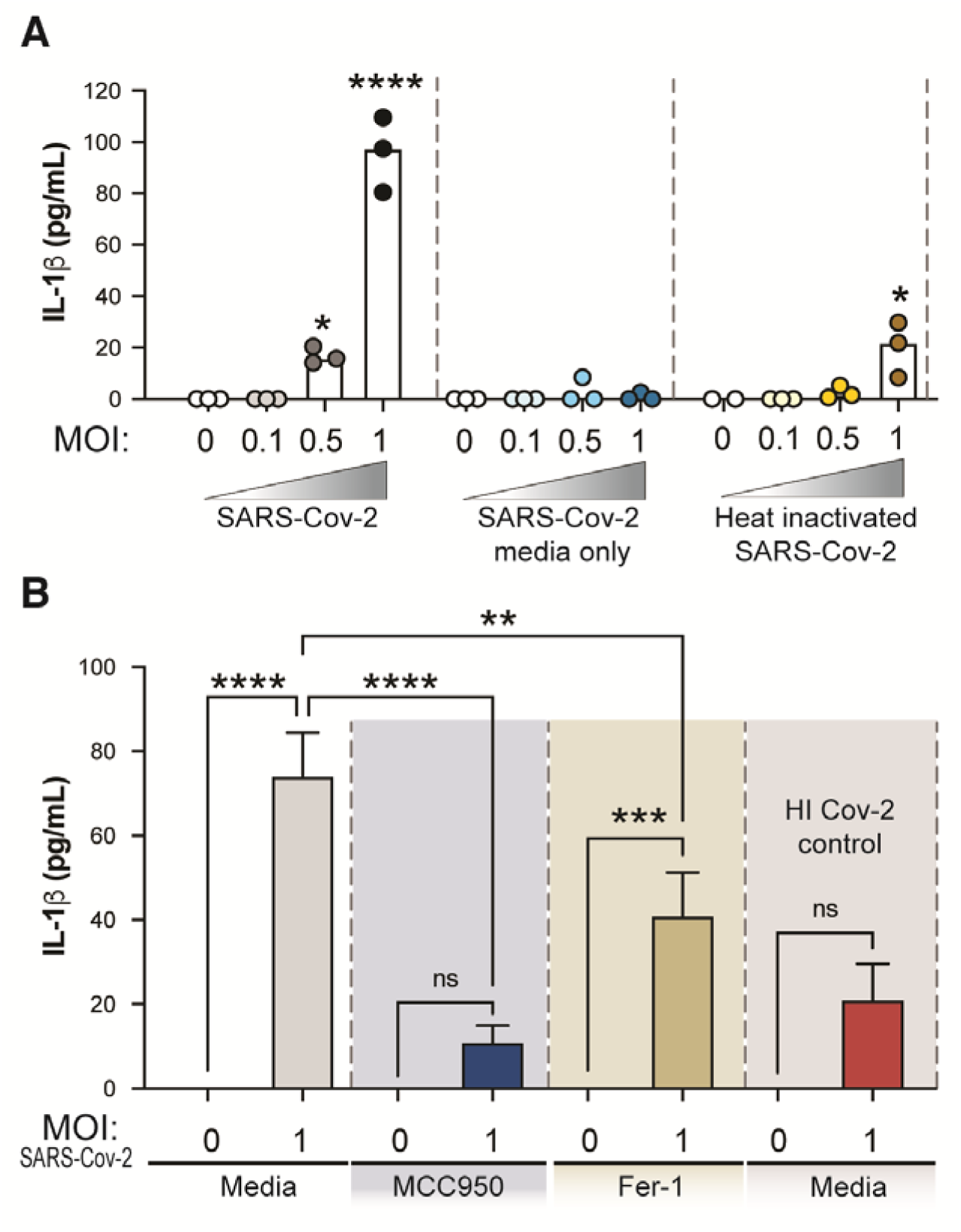
Exposure of human healthy control monocytes to SARS-CoV-2 *in vitro* induces IL-1β by a mechanism involving both inflammasome activation and lipid peroxidation. (**A**) Elutriated monocytes isolated from fresh healthy donors PBMCs were co-cultured with live SARS-CoV-2 (USA- WA1/2020) at distinct MOIs (0.1, 0.5 or 1), media only (mock) or heat inactivated SARS-CoV-2 (HI Cov-2) and IL-1β levels were measured in the culture supernatants after 24 hours by ELISA. Data represent the means ± SEM of triplicate samples. Statistical significance was assessed by one-way ANOVA analysis for the indicated experimental conditions. *P <0.05, ****P <0.0001. (**B**) Healthy donor monocytes were incubated with SARS-CoV-2 at MOI of 1 and treated or not with the NLRP3 inhibitor, MCC950 (10μM) or the lipid peroxidation inhibitor, Fer-1 (10μM). IL-1β levels were measured in culture supernatants after 24 hours by ELISA. Data represent the means ± SEM of 2 independent experiments pooled with 5-6 replicates. Statistical significance was assessed by one-way ANOVA analysis for the indicated experimental conditions. **P < 0.01, ***P <0.001, ****P <0.0001.

We then examined the direct role of NLRP3 and ROS generation on inflammasome activation in response to SARS-CoV-2 in our *in vitro* system, by incubating monocytes with NLRP3 inhibitor MCC950 or Ferrostatin-1 (Fer- 1), a compound known to inhibit lipid peroxidation. Interestingly, we observed that live SARS-CoV-2-induced IL-1β production was completely abrogated by MCC950 and partially inhibited by lipid peroxidation (Figure 6B). These *in vitro* results suggest that lipid peroxidation, a surrogate of oxidative stress, can further exacerbate NLRP3-inflammasome activation in response to SARS-CoV-2.

### Excessive oxidative stress response and inflammatory profile persist in COVID-19 patients after short-term recovery

It has been reported that some COVID-19 patients display lingering symptoms after disease recovery. This clinical condition known as post-acute COVID-19 syndrome is not yet well understood and has been described even in patients who experienced mild disease (Nalbandian et al., 2021; Yong, 2021). Since cell stress and inflammatory measures were elevated in COVID-19 patients, we sought to investigate whether this exacerbated inflammatory status persisted after a short recovery (approximately 52 days after infection onset). To do so, we first evaluated the frequency of monocytes along with expression of phenotypic or inflammatory markers such as CCR2, HLA-DR, ASC-speck formation, caspase-1 activity, mitochondrial superoxide generation, lipid peroxidation as well as cytokine production including IL-18, IL-1β, IL-6 and TNF-α (Figure 7A-L), by comparing those markers across recovered individuals and HCs. Among all parameters tested, we observed that CCR2 expression (Figure 7B), FLICA^+^ASC-speck monocyte frequency (Figure 7D), caspase-1 activity (Figure 7E), lipid peroxidation (Figure 7F), intracellular GSH levels (Figure 7H) and spontaneous cytokine release *in vitro* were all significantly altered in the COVID-19 recovered group *versus* HCs, indicating that short-term recovery does not afford a return to previous levels for these measurements (Figure 7). In contrast, longitudinal analysis of revealed a significant increase in CD14^low^CD16^+^ monocytes after recovery, suggesting a restoration of the normal distribution of monocytes subsets in peripheral blood (Figure 7A). Additionally, measurements associated with metabolic shift (Supplemental Figure 3C), cell activation (Supplemental Figure 4C-D), inflammasome activation (Supplemental Figure 4E-F), oxidative stress (Supplemental Figure 4G-H), as well as pro-inflammatory cytokine production (Supplemental Figure 4I-L) showed no significant difference between the acute and recovery phase. Importantly, the sustained cellular stress and inflammatory status found in the COVID-19 patients post-recovery was not associated with initial disease severity, since this inflammatory profile was observed regardless of patient classification at the acute phase of the disease as inpatients (closed symbols) or outpatients (open symbols) (Figure 7 and Supplemental Figure 4). These findings suggest that COVID-19-related stress and inflammatory responses persist after short-term recovery even in patients displaying mild symptoms at the acute phase of disease.

**Figure 7.**
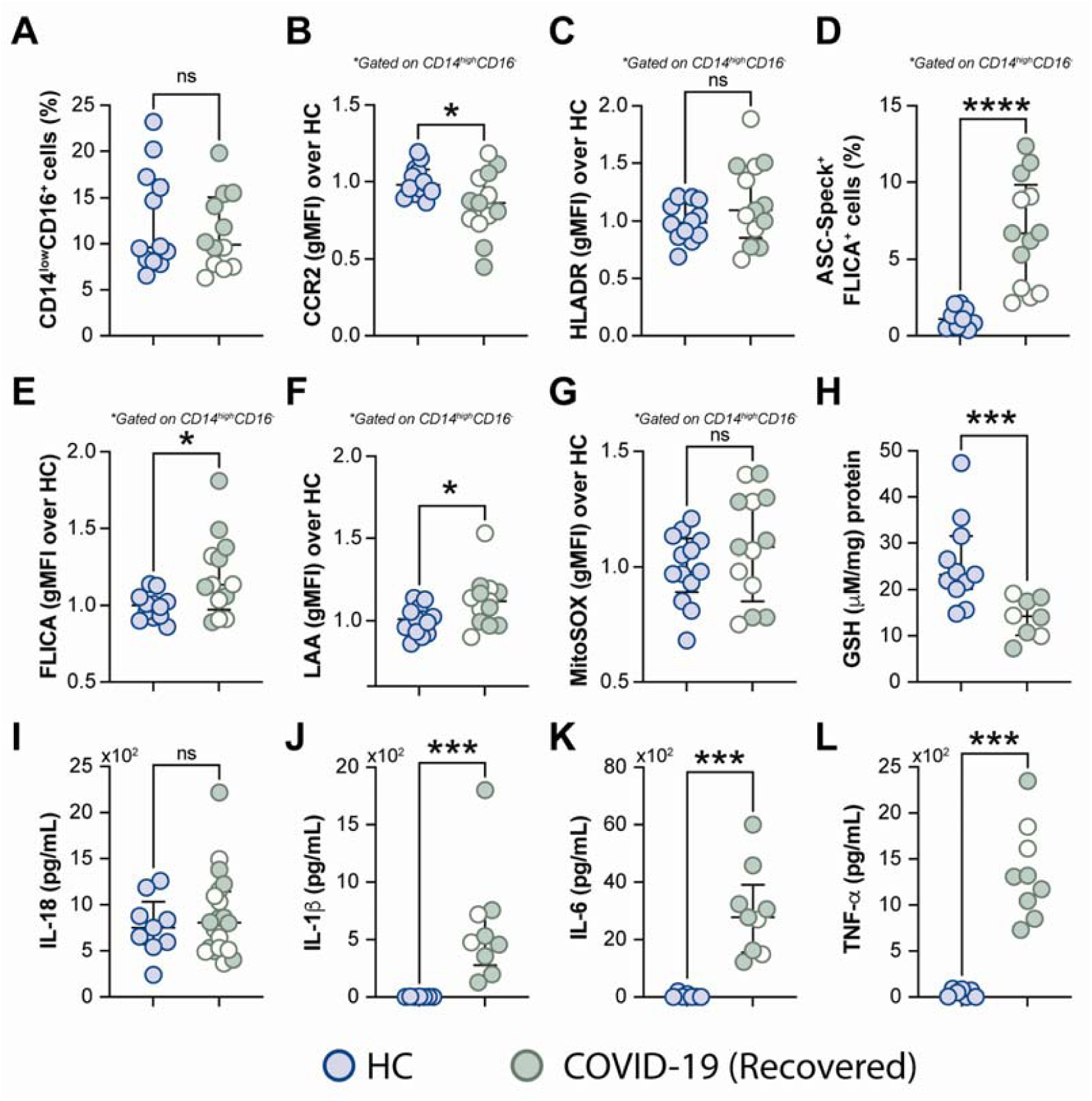
Measurements associated with inflammatory responses persist in COVID-19 patients after short-term recovery. Oxidative stress and inflammatory markers were compared between HCs and COVID-19 patients after recovery (median of 52 days IQR: 47.3-75.3, after infection onset) as follows: (**A**) Percentage of patrolling CD14^low^CD16^+^ monocyte subset, (**B**) CCR2 expression on monocytes, (**C**) HLADR expression on monocytes, (**D**) Percentage of ASC-speck^+^ CD14^high^CD16^-^ cells, (**E**) caspase-1 activity measured by FLICA staining, (**F**) lipid peroxidation (LAA) and (**G**) mitochondrial superoxide (MitoSOX) in classical monocytes, (**H**) intracellular glutathione levels in PBMC. (**I**) IL-18 levels were measured in plasma samples. (**J**-**L**) Spontaneous *in vitro* IL-1β, IL-6 and TNFα production by PBMC, respectively. Lines represent median values and interquartile ranges. Closed symbols indicate hospitalized patients (inpatients) and open symbols denote mild disease outpatients. Data were analyzed using the Mann-Whitney test. *P <0.05, ***P <0.01, ****P <0.01; *ns*, not significant.

## DISCUSSION

In SARS-CoV-2 infection, inflammatory monocyte-derived macrophages are the prominent immune cells in the infected lungs, playing a pivotal role as a major source of inflammatory mediators and thus significantly contributing to disease progression (Franks et al., 2003; Nicholls et al., 2003). Herein, we expand this knowledge by showing that approximately 10% of circulating CD14^high^CD16^-^ inflammatory monocytes from COVID-19 patients demonstrate ASC/Caspase-1/4/5 inflammasome complex formation, which appears to persist after short-term recovery. These aggregates are required for caspase-1-mediated IL-1 and IL-18 secretion, highlighting the role of CD14^high^CD16^-^ monocytes in COVID-19 pathogenesis. The enhanced caspase-1 activity of the pro-inflammatory monocytes strongly correlated with dysregulated oxidative stress status evidenced by mitochondrial dysfunction, mitochondrial superoxide generation and lipid peroxidation. Such clinical findings were confirmed *in vitro*, with oxidative stress identified to be partially involved in NLRP3-mediated IL-1β secretion by monocyte cultures exposed to SARS-CoV-2. Importantly, elevated inflammasome activation and aberrant oxidative stress response were both strongly correlated with worse clinical disease outcome, supporting the concept that myeloid-derived inflammatory responses are closely associated with disease severity in COVID-19.

Marked phenotypic differences were observed in the monocytic compartment in COVID-19 patients, including a striking depletion of the CD14^low^CD16^+^ patrolling monocyte subset, an expansion of the CD14^high^CD16^-^ classical/inflammatory monocytes and downmodulation of HLA-DR and CCR2 markers in the CD14^high^CD16^-^ cells as reported in other studies. Robust HLA-DR downregulation in monocytes from COVID-19 hospitalized patients compared to HCs has been previously shown by single- cell RNA sequencing analysis (Wilk et al., 2020). Dimensionality reduction analysis of monocytes also indicated a strong phenotypic shift towards the CD14^+^ monocyte subset in patients with acute respiratory distress syndrome (Wilk et al., 2020). More recently, the SARS-CoV-2 protein ORF7a has been shown to interact with CD14^+^ monocytes, triggering downmodulation of HLA- DR and production of proinflammatory cytokines *in vitro* (Zhou et al., 2021). It is not clear, however, whether specific viral factors may also contribute to the downregulation of CCR2 on classical monocytes. CCR2 expression on classical CD14^high^CD16^-^ monocytes is known to mediate bone marrow egress and recruitment into inflamed tissues in response to CCL2 (Phillips et al., 2005). Several studies have shown elevated levels of CCL2 along with other chemokines in serum and bronchoalveolar fluid (BALF) samples obtained from COVID-19 patients, which may contribute to classical monocyte expansion, substantial mononuclear cell infiltration into the lungs, and consequently tissue damage (Khalil et al., 2021; Merad and Martin, 2020; Wilk et al., 2020). Thus, our data suggest downmodulation of CCR2 as a possible compensatory mechanism of CD14^high^ monocytes in response to sustained overproduction of CCL2 or continuous trafficking of CCR2^+^ cell into tissues (Aragay et al., 1998). Intriguingly, our finding showing a failure of CCR2 expression to return to normal levels after short-term recovery indicates persistent chemokine overexpression and monocyte recruitment even after the acute phase of the infection.

Our results also indicate NLRP3 as the main inflammasome sensor promoting the poor outcome of COVID-19, in agreement with recently reported studies (Marchetti et al., 2021; Rodrigues et al., 2021). In this regard, the presence of NLRP3 inflammasome aggregates has been previously described in PBMCs and autopsy tissue samples from COVID-19 patients (Rodrigues et al., 2021; Toldo et al., 2021). Our findings extend these observations by revealing that aberrant oxidative stress responses evidenced by mitochondrial superoxide production and lipid peroxidation strongly associate with caspase-1 activity, suggesting a crosstalk between oxidative stress and inflammasome activation in SARS-CoV-2 infection. In fact, intracellular ROS production has been suggested as a key trigger of NLRP3 inflammasome activation as a result of either direct pathogen sensing or in response to DAMPs released from dying cells (Martinon, 2010; Tschopp and Schroder, 2010). Previous studies have reported that SARS-CoV transmembrane pore-forming protein, known as viroporin SARS-CoV 3a or ORF3a, activates the NLRP3 inflammasome complex in LPS-primed macrophages through a mechanism dependent on K+ efflux and mitochondrial ROS (Chen et al., 2019). In addition, ORF3a was also shown to promote inflammasome assembly through TRAF3-mediated ubiquitination of ASC, while another viral protein, ORF8b seems to interact directly with the leucine-rich repeat domain of NLRP3 to stimulate its activation independent of ion channel activity (Shi et al., 2019). More recently, an *in vitro* study showed that SARS-CoV-2 viroporin protein induces the NLRP3-inflammasome formation in a lung epithelial cell line (Xu et al., 2020), suggesting that SARS-CoV-2 may trigger ROS-mediated NLRP3-inflammasome activation similarly to SARS-Cov.

It remains controversial whether circulating blood monocytes can express the receptor ACE-2, the dominant entry receptor for SARS-CoV-2, and thus be actively infected (Gheblawi et al., 2020; Zamorano Cuervo and Grandvaux, 2020). Although initial work failed to identify ACE-2 on most tissue-resident macrophages and monocytes (Merad and Martin, 2020), other studies have shown ACE-2 protein expression in human circulating monocytes and macrophages by immunoblotting or flow cytometry (Rutkowska-Zapala et al., 2015; Zhang et al., 2021). More recent reports suggest that SARS-CoV-2 can actively infect primary human monocytes, resulting in NLRP3 activation and consequently IL-1β secretion (Ferreira et al., 2021; Junqueira et al., 2021; Rodrigues et al., 2021). In the present study we were unable to observe productive infection in human monocytes exposed to the SARS-CoV-2 virus *in vitro* (Supplemental Figure 5). A possible explanation for the discrepancy between our results and others relates to the different approaches used to evaluate SARS-CoV-2-monocyte infection. In this regard, here we evaluated productive SARS-CoV-2 infection by exposing VERO E6 indicator cells which are known to be virus susceptible, to monocyte lysates, whereas other studies have employed rt-PCR or fluorescent-protein reporter virus to evaluate SARS-CoV-2 infection in human monocyte cultures.

Nevertheless, in agreement with a role for direct monocyte sensing of SARS-CoV-2 in NLRP3 activation, we observed that exposure of primary human HC monocytes to the active virus was sufficient to promote NLRP3- driven IL-1β production. Importantly, SARS-CoV-2-induced IL-1β secretion was partially suppressed by treating cells with a lipid peroxidation inhibitor, Fer-1. This observation is consistent with a recent report showing that monocyte-derived macrophages and dendritic cells can be activated by SARS-CoV-2 to induce both cytokine production and cell death even in the absence of productive infection (Zheng et al., 2021). Our findings, however, do not rule out the possibility that monocytes/macrophages are able to be productively infected in patients. In addition to ACE2 binding, other potential routes of SARS-CoV-2 entry in monocytes have also been proposed such as binding of the spike protein to the CD147 receptor, antibody-mediated phagocytosis via Fc gamma receptors, as well as engulfment of dying infected cells, (Bournazos et al., 2020; Junqueira et al., 2021; Wang et al., 2020). Nonetheless, further studies with SARS-CoV-2 variants displaying different mutations are also needed to investigate whether changes in the viral genome may affect viral uptake by monocytes/macrophages.

A major observation of the present study is that the inflammatory profile of circulating monocytes persists in COVID-19 patients, post-recovery regardless of their disease severity during acute infection (Figure 7 and Supplemental Figure 4). It has been reported that about 80% of hospitalized patients with COVID-19 displayed at least one long-term symptom for several months after discharge, particularly fatigue and dyspnea (Arnold et al., 2020; Carfi et al., 2020). Moreover, lingering symptoms and functional impairment up to eight months after contracting mild SARS-CoV-2 infection was also reported in a group of low-risk healthcare workers (Havervall et al., 2021). Indeed, some patients in our cohort, who had experienced either mild or severe symptoms during acute COVID-19, have also reported the persistence of at least one of the current known COVID-19 symptoms following recovery from infection. However, we were unable to establish a close association between the biomarkers tested in this study and any of the residual symptoms post-recovery. The pathophysiology, risk factors and treatment of post-acute COVID-19 is currently poorly understood. In that sense, our observations of sustained dysregulated oxidative stress and inflammasome activation in monocytes after short-term recovery support the current hypothesis that long term COVID-19 is driven by persistent pathological inflammation and suggest the pathways involved as potential targets for therapeutic intervention (Nalbandian et al., 2021; Yong, 2021). In this regard, it will be important to evaluate whether persistent immune activation after COVID-19 recovery relies on the presence of undetectable SARS-CoV-2 virus, viral RNA and/or viral antigens as well as assess the contribution of unrepairable cellular and tissue damage.

The limitations of the current study include lack of critically ill ventilated ICU patients (as participants had to be able to provide consent), lack of multiple longitudinal sampling and small sample size. Nevertheless, our findings collectively corroborate the hypothesis that early exposure of alveolar macrophages and endothelial cells to SARS-CoV-2 may induce oxidative stress dysregulation and aberrant cytokine secretion. This initial cell priming event may contribute to the enhanced recruitment to the site of infection of pre-activated circulating monocytes pre-committed to inflammasome activation and aberrant oxidative stress response, resulting in exacerbation of tissue immunopathology and ultimately organ failure. In this context, recent advances in targeting ROS by administration of glutathione or glutathione precursors (N-acetyl-cysteine, NAC), as well as IL-1 signaling by administrating Anakinra, a IL-1 Receptor antagonist (IL-1Ra), have been reported to prevent inflammation and/or worse clinical outcome in COVID-19 patients (Cauchois et al., 2020; Cavalli et al., 2020; Horowitz et al., 2020; Huet et al., 2020; Ibrahim et al., 2020; Kyriazopoulou et al., 2021a; Kyriazopoulou et al., 2021b). Thus, our findings suggest that early treatment with antioxidants and IL-1 signaling inhibitors may represent potential therapeutic intervention for COVID-19 by preventing deleterious effects downstream of the inflammasome pathway and consequently tissue immunopathology and that such interventions may have a role in the recovery phase of the disease.

## Materials and methods

### Plasma biomarker measurements

Cryopreserved plasma samples from HCs and patients were assessed to measure levels of the following inflammatory biomarkers according to the manufacturer’s instructions. Levels of IFN-γ, IL-2, IL-6, IL-7, IL-8, IL-10, IL-12p70, TNF-α, IL-27, E-Selectin, P-Selectin, ICAM-3, SAA, CRP, VCAM-1, ICAM-1 and IL-18 were quantified using electrochemiluminesence kits from Meso SCALE (Gaithersburg, MD, USA). Levels of ferritin heavy chain (ferritin-H) were determined by enzyme-linked immunosorbent assay (ELISA) from Thermo Fisher Scientific (Rockford, IL, USA). D-dimer levels were measured using Enzyme Linked Fluorescence Antibody kits from BioMerieux, MA, USA. Levels of iron was assessed using a commercial kit from Novus Biologicals (Centennial, CO). Concentrations of catalase, superoxide dismutase activity (SOD) and total oxidant status were measured using kits from Sigma-Aldrich (St. Louis, MO, USA). Levels of sCD14 and sIL-6Ra were quantified by ELISA from R&D Systems (Minneapolis, MN, USA). Lipid peroxidation in plasma was assessed using an assay kit from Cayman Chemical, which measures the formation of malondialdehyde (MDA).

### Cell culture and treatments

Cryopreserved ficoll-isolated peripheral blood mononuclear cells (PBMCs) from patients or healthy control individuals (HCs) were thawed and resuspended in RPMI-1640 media (Corning, NY, USA) supplemented with 10% heat-inactivated human AB serum (Gemini Bio-Products, West Sacramento, USA) and 0.05% benzonase (MilliporeSigma, USA). Cells were rested for 1 hour at 37 °C and 5% CO_2_ and subsequently plated at 10^6^ cells/well in round bottom 96-well plates (Corning Costar^TM^, MilliporeSigma, USA) followed by immune staining. Alternatively, cells were incubated for 3hr, in the presence or absence of MCC950 (3 μM; Invivogen, CA, USA) or colchicine (10 μM; Sigma-Aldrich, St Louis, MO, USA) for inflammasome inhibition. IL-1β, IL-6 and TNF-α were quantified in culture supernatants by using Multi-Analyte Flow Assay Kit (LEGENDplex) from Biolegend, following the manufacturer’s instructions.

### Inflammasome and mitochondrial status assessment by Imaging Flow Cytometry

Inflammasome complex assembly was evaluated by detection of ASC speck formation by imaging flow cytometry, as previously described (Lage et al., 2019). Briefly, PBMCs were incubated with the fluorochrome inhibitor of caspase-1/4/5 (FAM-FLICA, Immunochemistry technologies (ICT), Bloomington, MN) following the manufacturer’s instructions, for 50 min at 37 °C, to allow for binding of FLICA to activated inflammatory caspases. Cells were washed twice with the FLICA kit wash buffer and then incubated with LIVE/DEAD Fixable AQUA Dead Cells Stain (Thermo Fisher, USA) for 15 min at RT, followed by extracellular staining in PBS + 1% BSA with the following fluorochrome-conjugated antibodies for monocyte phenotyping: anti-CD14 BV605 (Clone:M5E2), anti-CD16 PE-Cy7 (Clone:3G8), and anti-CD3 PE (Clone:HIT3a) from BioLegend, San Diego, CA, USA; anti-CD20 PE (Clone:2H7), anti-CD19 PE (Clone:SJ25C1), anti-CD2 PE (Clone:RPA-2.10), from eBioscience, San Diego, CA; anti-CD56 PE [(Clone: B159 (RUO)], anti- HLA-DR BV421 [Clone: G46-6 (RUO)] and anti-CD66b (Clone:G10F5) from BD Biosciences, Franklin Lakes, New Jersey. Cells were fixed and permeabilized with Cytofix/Cytoperm (BD Biosciences, USA) overnight at 4°C and stained for 1h at RT for intracellular ASC, with anti-ASC/TMS1 AF647 antibody from Novus Biologicals, Littleton, CO. In parallel, cells were stained with monocyte markers and 200 nM Mitotracker Red (MitoTracker™ Red CMXRos, Invitrogen) followed by ASC intracellular staining for mitochondrial membrane potential evaluation. Cells were acquired using a 12-channel Amnis ImageStreamX Mark II (MilliporeSigma) imaging flow cytometer and the integrated software INSPIRE (MilliporeSigma) was used for data collection. Images were analyzed using image-based algorithms in the ImageStream Data Exploration and Analysis Software (IDEAS 6.2.64.0, MilliporeSigma) as described previously (Lage et al., 2019).

### Flow Cytometry

PBMCs were stained with the following list of fluorescently labeled antibodies against cell surface markers subsequently detected by flow cytometry: anti- CD14 BV605 (Clone:M5E2), anti-CD16 PE-Cy7/BV711 (Clone:3G8), and anti- CD3 PE (Clone:HIT3a) from BioLegend; anti-CD20 e450 (Clone:2H7), anti- CD19 e450 (Clone:SJ25C1), anti-CD2 e450 (Clone:RPA-2.10), from eBioscience; anti-CD56 e450 [(Clone: B159 (RUO)], anti-HLA-DR APC-Cy7 [Clone: G46-6 (RUO)] and anti-CD66b e450 (Clone:G10F5) and CCR2 PerCP Cy5.5 [Clone:] from BD Biosciences. Staining protocol proceeded as described above for imaging flow cytometry analysis. Surface Glut-1 receptor expression was detected by binding to the Glut-1 ligand fused to enhanced green fluorescent protein (GFP) (Metafora Biosystems, Evry, France), as previously described (Hammoud et al., 2019). In brief, 10^6^ cells were incubated for 20 minutes with 10 µL of Glut-1–GFP solution prior to Live/Dead staining using LIVE/DEAD Cell Viability Assay (Life Technologies, CA, USA). GFP fluorescence was detected by staining cells with Alexa Fluor 647 anti- GFP antibody at 1:200 (Biolegend). Data were acquired on a BD Fortessa flow cytometer (BD Biosciences). All compensation and gating analyses were performed using FlowJo 10.5.3 (TreeStar, Ashland, OR, USA).

### Lipid peroxidation assay

Lipid peroxidation in total PBMC cultures was assessed by using the Click-iT Lipid Peroxidation Imaging Kit (Life Technologies, SA, USA) according to the manufacturer’s instructions. Cells were incubated with the LAA reagent (alkyne-modified linoleic acid) for detection of lipid peroxidation-derived protein modifications at 37°C for 1 h, and then washed with 1× DPBS. After cell centrifugation (1,500 rpm for 5 min) to remove the LAA reagent, cells were incubated with antibodies to measure lipid peroxidation only in CD14+ monocytes. Live/Dead staining was performed as described previously above and the cells fixed by adding cytofix/cytoperm (BD bioscience, USA). After completing the LAA staining, fixed cells were washed, and resuspended in 1× DPBS, and then LAA fluorescence was analyzed by flow cytometry.

### Measurement of intracellular GSH levels

Intracellular GSH levels were measured in PBMC lysates by using a Glutathione Assay Kit (Cayman Chemical, USA) according to the manufacturer’s instructions. Total GSH levels were normalized by total protein concentration determined by using a Pierce BCA protein Assay kit (Thermo Fisher, IL, USA).

### Mitochondrial superoxide assay

Mitochondrial superoxide was detected by using a flow cytometry–based assay. Total PBMCs were washed with Hanko calcium and magnesium (HBSS/Ca/Mg, Gibco, USA) to remove residual media by centrifugation. Cell pellet was stained with MitoSOX probe (Life Technologies, USA) at 37°C for 30 min following the manufacturer’s protocol. Cells were next washed with HBSS/Ca/Mg and incubated with antibodies to analyze CD14^+^ monocytes by flow cytometry. Cells fixed with 2% paraformaldehyde at room temperature for 30 min. Fluorescence intensity of the probe was then measured by flow cytometric analysis.

### *In vitro* exposure of human monocytes to SARS-CoV-2

Elutriated monocytes isolated from fresh PBMCs were co-cultured with SARS-CoV-2 USA-WA1/2020 at different MOIs for 3 hours at 37°C under low serum conditions (2% fetal calf serum) in a Biosafety Level 3 facility. Control cells were co-cultured with heat-inactivated SARS-CoV-2 USA-WA1/2020 at a MOI of 1, mock viral supernatant at a MOI of 1 or media alone. After 3 hours of viral exposure, monocytes were treated with Ferrostatin-1 (Fer-1) (10`μ`M; Selleck, USA), MCC950 (10µM; Invivogen, CA, USA) or vehicle (DMSO; Sigma-Aldrich, St. Louis, MO, USA) and thus incubated for an additional 21 hours at 37°C. Cell supernatants were harvested for quantification of IL-1β by ELISA following the manufacturer’s directions (Sigma-Aldrich, USA).

To determine viral titers by TCID50 assay, cells were harvested, lysed by freeze/thawing and plated in triplicate onto Vero E6 cells using 10-fold serial dilutions. Viral stock was used as a positive control. Plates were stained with crystal violet after 96 hours to assess CPE. Viral titers were determined using the Reed-Muench method.

### Statistical Analyses

Statistical analyses were performed using non-parametric Mann-Whitney or Kruskal-Wallis test in GraphPad Prism 8.0.1 software (GraphPad, USA). Data are presented as median with interquartile ranges. Spearman’s correlation analyses were performed using R. Differences between groups or parameters were considered significant when *p* < 0.05.

### Study participants and approval

Study participants were enrolled in the NIH clinical protocol: COVID-19- associated Lymphopenia Pathogenesis Study in Blood (CALYPSO), NCT04401436, which recruited at the NIH clinical center, Washington Hospital Center and Georgetown University Hospital. Healthy volunteer blood samples were obtained under the protocol 99-CC-0168 and were de-identified prior to distribution. Protocols in this study were reviewed and approved by the National Institutes of Health (NIH) Central Intramural Institutional Review

Board (IRB). All participants provided written informed consent prior to any study procedures in accordance with the Declaration of Helsinki. Patients enrolled in this study were classified according to their highest oxygen requirement up until the time of the research blood draw in (1) mild cases, in which patients required 4 Liters or less, (2) moderate cases, when their requirement was up to 50% oxygen concentration (FiO2), and (3) severe cases, in which patients required over 50% FiO2 or ICU care. Only patients who were able to consent themselves were eligible to participate, which limited the enrollment of critically ill ventilated ICU patients. In addition, 14 patients were sampled longitudinally, at the acute phase of the disease and after a short-term recovery period (approximately 52 days after infection onset) and 2 patients were enrolled after their recovery.

## AUTHOR CONTRIBUTIONS

SLL, EPA, AS and IS were involved in conception and design of the study and experiments. SLL, EPA, KH, AR, SN and JR performed experiments and/or data analysis. EL, PK, GW, RP, FG, AK, JR, MM, AL and IS recruited patients, assisted in clinical care and helped with data curation and/or visualization. AR, AS, IS, JPS and HDH provided resources and reagents. SLL, EPA and IS were involved in drafting the manuscript. All authors have reviewed and approved the final manuscript.

## Data Availability

I state that all data are available in the manuscript files.

## ACKNOWLEDGMENTS

This project was supported in part by the intramural research program of NIAID/NIH, with federal funds from the National Cancer Institute, National Institutes of Health, under Contract No. HHSN261200800001E. KH was supported by a Malahan Institute Postdoctoral Fellowship. The content of this publication does not necessarily reflect the views or policies of the Department of Health and Human Services, nor does mention of trade names, commercial products, or organizations imply endorsement by the U.S. Government. The authors declare no competing financial interests. We are grateful to the patients enrolled in protocols and blood bank healthy donors for making this study possible. We also thank Drs Kumar and Woltmann and their associated staff at WHC and Georgetown, as well as the OP8 Infectious Diseases Clinic staff for their assistance in patient recruitment and evaluation. We also thank David Stephany for his technical support.

## Notes

### Competing Interest Statement

The authors have declared no competing interest.

### Author Declarations

Study participants were enrolled in the NIH clinical protocol: COVID-19-associated Lymphopenia Pathogenesis Study in Blood (CALYPSO), NCT04401436, which recruited at the NIH clinical center, Washington Hospital Center and Georgetown University Hospital. Healthy volunteer blood samples were obtained under the protocol 99-CC-0168 and were de-identified prior to distribution. Protocols in this study were reviewed and approved by the National Institutes of Health (NIH) Central Intramural Institutional Review Board (IRB). All participants provided written informed consent prior to any study procedures in accordance with the Declaration of Helsinki.

