## Supplemental Material for "Persistent oxidative stress and inflammasome activation in CD14^high^CD16^-^ monocytes from COVID-19 patients"

**Supplemental Figures**

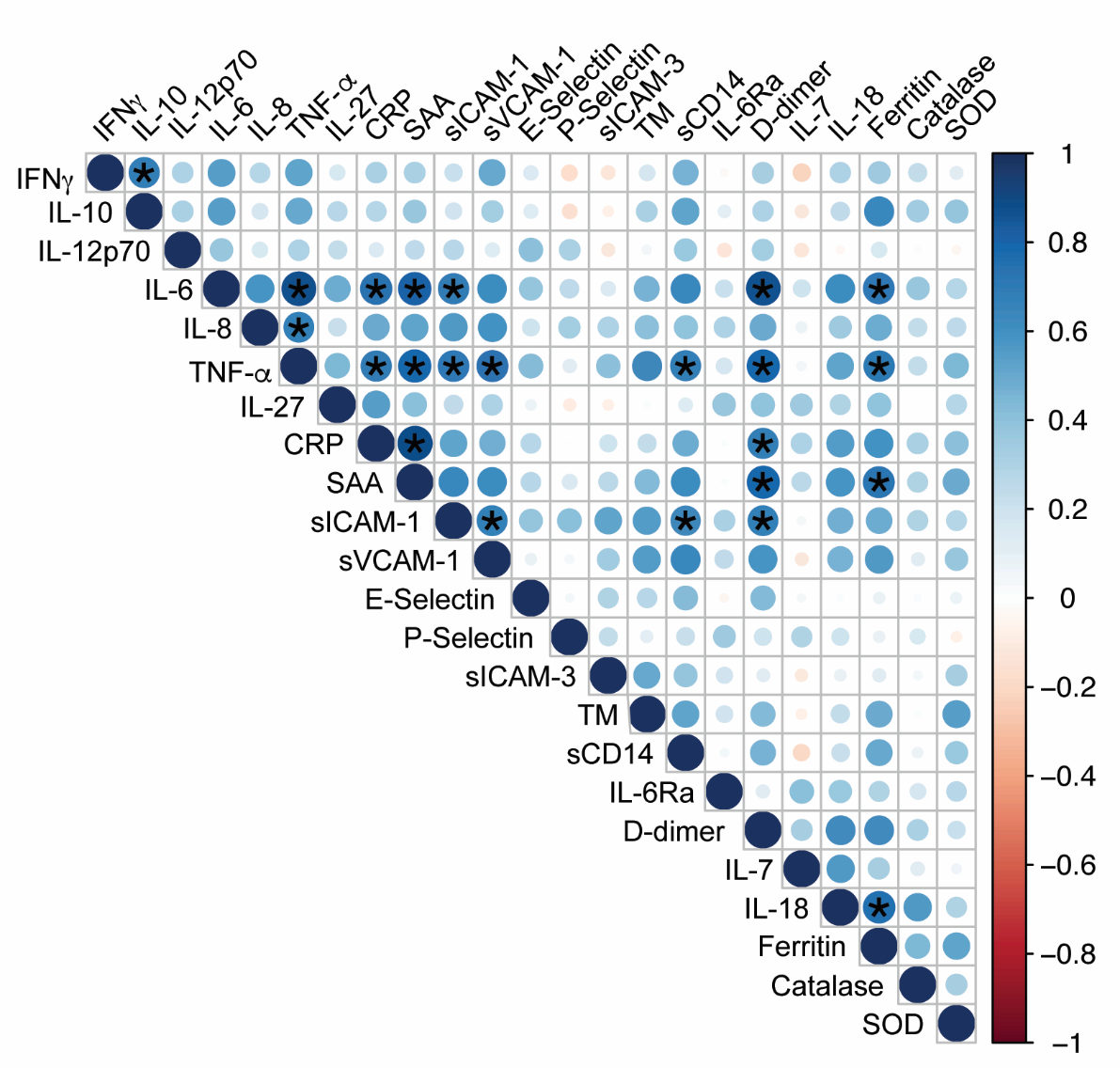

**Supplemental Figure 1. Correlation of inflammatory response in the plasma of COVID-19 patients.** Multi-parameter Spearman’s analysis was performed to evaluate the correlation of several markers associated with inflammatory response in plasma from COVID-19 patients (*P <0.05 or less following Holm-Benforroni test for multiple comparisons).

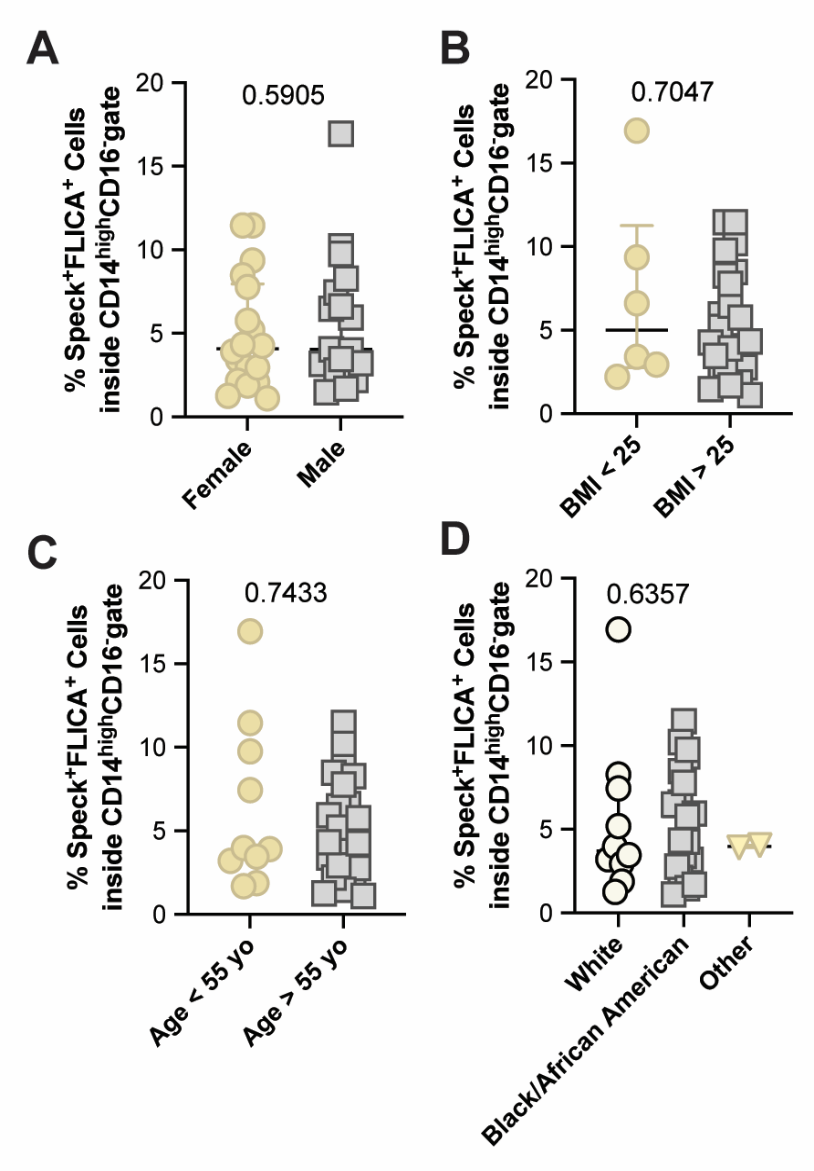

**Supplemental Figure 2. Inflammasome complex formation is not associated with underlying demographic characteristics of COVID-19 patients.** Percentages of speck^+^FLICA^+^ cells within the classical CD14^high^CD16^-^ monocyte compartment were compared between female *versus* male COVID-19 patients in (**A**), patients with low (<25) or high (>25) BMI in (**B**), patients that were <55yo versus > 55yo in (**C**) and patients within distinct race/ethnicity backgrounds in (**D**). Lines represent median values and interquartile ranges. Data were analyzed using the Mann-Whitney test.

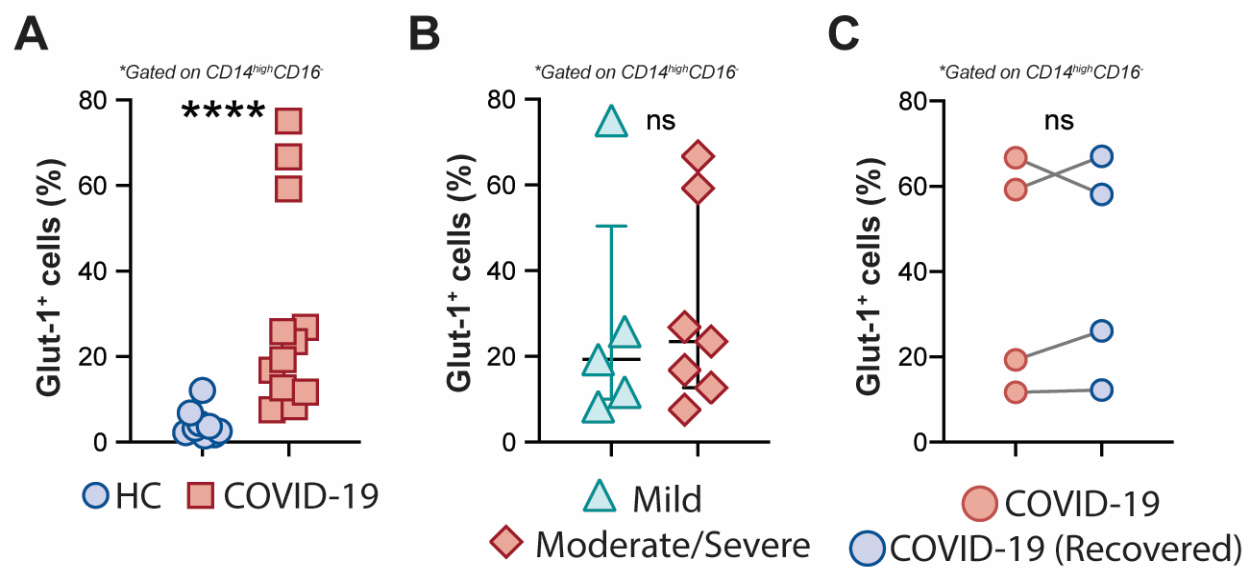

**Supplemental Figure 3. COVID-19 patients monocytes express increased levels of the glucose receptor-1.** Percentage of CD14^high^CD16^−^ monocytes expressing the Glut-1 receptor was compared between healthy controls (HC, n=9) and COVID-19 patients (n=12) in (**A**) or between COVID-19 patients experiencing mild *versus* moderate-severe disease in (**B**). Data are presented as median with interquartile range. ****P < 0.001, when Mann-Whitney test was applied. (**C**) Paired analysis between COVID-19 patients during the acute phase of the disease and same individuals approximately 1 and a half month after disease onset (recovered) were done comparing the percentage of CD14^high^CD16^−^ monocytes expressing the Glut-1 receptor. Data were analyzed using the Wilcoxon test. *ns*, not significant.

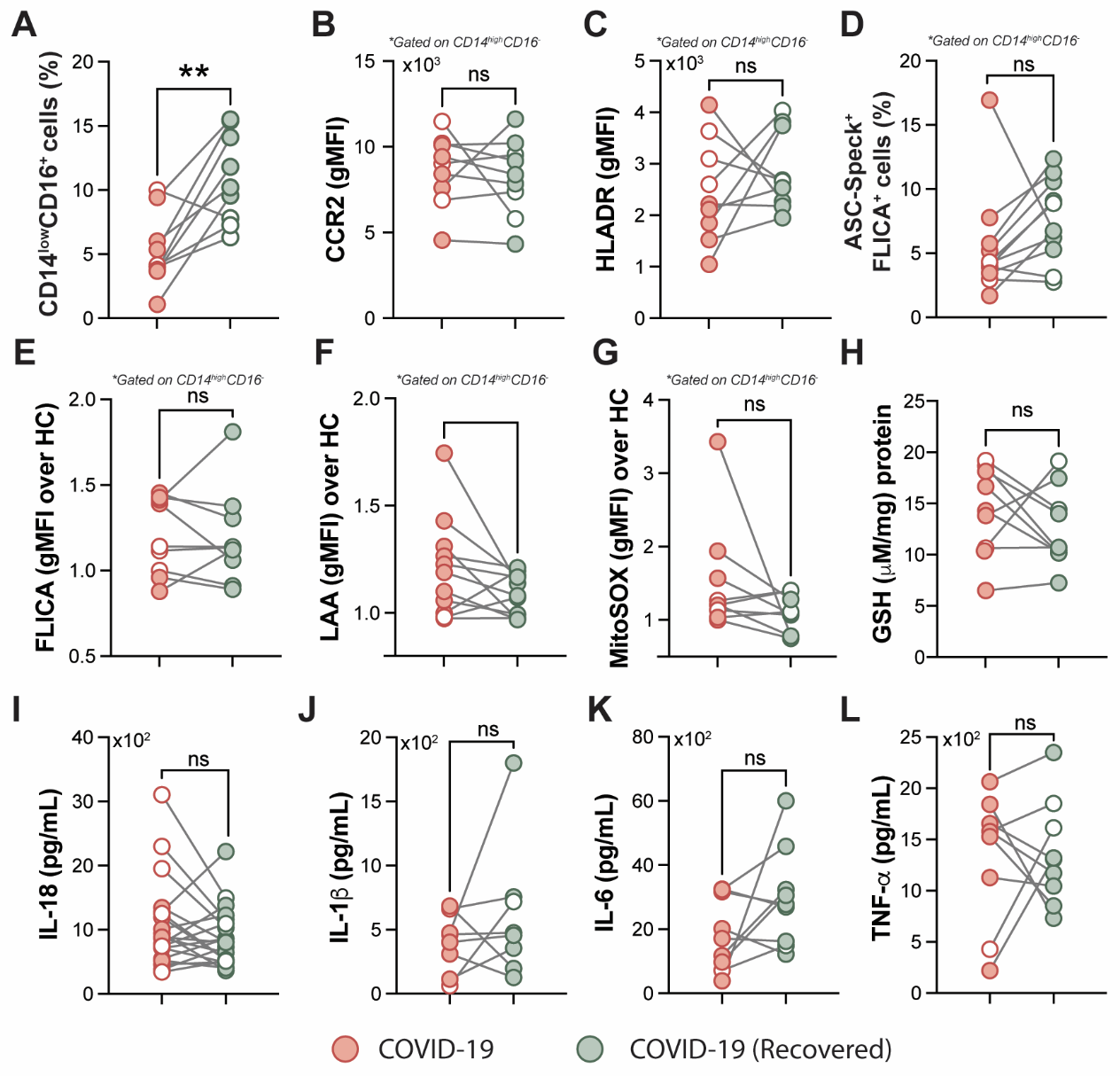

**Supplemental Figure 4. COVID-19 patients display persistent elevated levels of oxidative stress and inflammation after recovery from acute infection.** Paired analysis between COVID-19 patients during the acute phase of the disease and same individuals at recovery (median of 52 days IQR: 47.3-75.3, after infection onset) were performed comparing various oxidative stress and inflammatory markers as follows: (**A**) Percentage of patrolling CD14^low^CD16^+^ monocyte subset, (**B**) CCR2 expression on monocytes, (**C**) HLADR expression on monocytes, (**D**) Percentage of ASC-speck^+^ CD14^high^CD16^-^ cells, (**E**) caspase-1 activity measured by FLICA staining, (**F**) lipid peroxidation (LAA) and (**G**) mitochondrial superoxide (MitoSOX) in classical monocytes, (**H**) intracellular glutathione levels in PBMC. (**I**) IL-18 levels were measured in plasma samples. (**J**-**L**) Spontaneous *in vitro* IL-1β, IL-6 and TNFα production by PBMC, respectively. Lines represent median values and interquartile ranges. Closed symbols indicate hospitalized patients (inpatients of various disease severity) and open symbols denote mild disease outpatients. Data were analyzed using the Wilcoxon test. **P <0.01; *ns*, not significant.

**Supplemental**
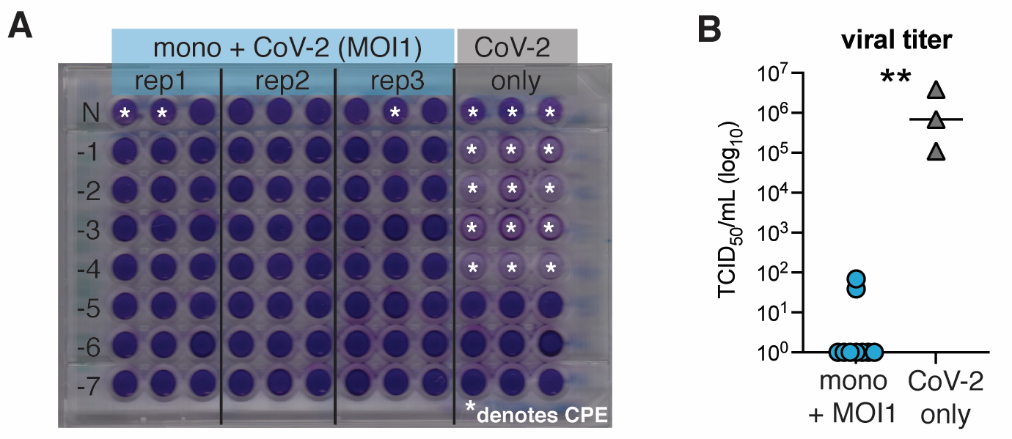
**Figure 5. Exposure of healthy donor-derived human monocytes to SARS-CoV-2 does not induce robust productive infection measured by the TCID50 assay.** Elutriated monocytes isolated from fresh healthy donor PBMCs were co-cultured with SARS-CoV-2 (USA-WA1/2020) at MOI of 1 for 24 hours. Cells were harvested, lysed and the TCID50 assay was employed to determine viral titers as described in Methods. Viral stock was used as a positive control. (**A**) Image of the TCID50 plate after Vero E6 cells exposure to monocyte lysates (mono+CoV-2) or viral stock (CoV-2 only) for 96 hours. (**B**) Quantification of viral load determined by TCID50 assay showed in A. **P <0.01 when Mann-Whitney test was applied. Representative data from two independent experiments in triplicate (means ± SEM).  Statistical significance was assessed by one-way ANOVA analysis for the indicated experimental conditions. **P < 0.01, ***P <0.001, ****P <0.0001.

**Supplemental Tables**

**Supplemental TABLE 1. Plasma Biomarkers of Mild *versus* Moderate/Severe groups (median values with IQR in parenthesis).**

| Biomarker (pg/ml) | Mild | Mod-Severe | p-value |
| --- | --- | --- | --- |
| N | 16 | 9 | NA |
| IFN-γ | 10.5 (5.3-26.4) | 6.74 (2.9-11.7) | 0.42 |
| IL-10 | 1.85 (0.67-3.16) | 0.67 (0.57-1.75) | 0.28 |
| IL-12p70 | 0.30 (0.17-1.57) | 0.26 (0.20-0.39) | 0.56 |
| IL-2 | 0.29 (0.14-0.43) | 0.21 (0.13-0.26) | 0.43 |
| IL-6 | 7.28 (1.52-16.6) | 10.3 (3.91-273) | 0.25 |
| IL-8 | 5.44 (2.93-8.33) | 4.88 (3.80-5.69) | 0.72 |
| TNF-α | 3.40 (2.11-5.77) | 3.72 (2.88-4.81) | 0.76 |
| IL-27 | 2898 (1712-4336) | 3295 (2255-5155 | 0.93 |
| CRP (mg/L) | 128.9 (29.0-235.1) | 97.0 (73.8-250.5) | 0.72 |
| SAA (mg/L) | 236.2 (22.8-468.1) | 369.3 (241.8-484.1) | 0.39 |
| sCD14 (mg/L) | 1.80 (1.46-2.48) | 1.83 (1.38-2.66) | 0.93 |
| IL-6Rα | 3851 (3407-4372) | 4535 (3874-5526) | 0.07 |
| D-dimer (ng/ml) | 1286 (585-2209) | 1886 (1571-3992) | 0.11 |
| Ferritin (ng/ml) | 1285 (204-1822) | 1288 (1021-1681) | 0.57 |

* **p-value** when Mann-Whitney was applied.
